# Investigating Data-Driven Climate-Adaptive Optimization of *P. vivax* Malaria Interventions in Seoul, the Republic of Korea

**DOI:** 10.64898/2025.12.12.25342125

**Authors:** Jiwon Han, Gerardo Chowell, Eunok Jung

## Abstract

*Plasmodium vivax* malaria control requires addressing unique challenges such as latent hypnozoite reservoirs, relapse-driven persistence, and strong climatic modulation of transmission. This study introduces a novel, integrative modeling–optimization framework that couples a relapse- and climate-aware transmission model with structural identifiablity analysis and metaheuristic optimization to design adaptive intervention strategies. Using surveillance data from Seoul, Korea, we calibrated key parameters through identifiability-guided estimation and optimized 48 monthly decision variables representing both pharmaceutical (tafenoquine substitution) and non-pharmaceutical interventions under resource constraints. The Improved Multi-Operator Differential Evolution (IMODE) algorithm efficiently navigated the high-dimensional, non-convex decision space, yielding climate-adaptive intervention schedules that reduced relapses by 80% and total infections by 16.6% relative to primaquine-only baselines. Economic analysis confirmed substantial cost-benefit (IBCR = 6.04), even under limited tafenoquine stockpiles. This work provides one of the first demonstrations of a structurally identifiable, climate-sensitive malaria model directly coupled to global evolutionary optimization, bridging mechanistic modeling with operational decision-making. The framework is broadly transferable to other vector-borne or climate-sensitive diseases, supporting data-informed elimination strategies in the face of environmental and logistical uncertainty.

**Highlights:**

- Climate-adaptive P. vivax model with identifiability-guided optimization
- Robust parameter calibration of relapse dynamics using Seoul data
- IMODE optimizes 48 monthly interventions under resource constraints
- 80% relapse reduction and strong cost-benefit (IBCR=6.04) achieved
- Transferable framework for climate-sensitive disease elimination planning

## 1. Introduction

Plasmodium vivax malaria presents unique epidemiological challenges due to its dormant liver-stage parasites (hypnozoites) that enable relapse infections months or years after initial treatment [1, 2]. Unlike *P. falciparum*, effective *P. vivax* control requires addressing both active infections and preventing relapses through radical cure protocols. Climate change further complicates control efforts by altering vector population dynamics and extending transmission seasons [3, 4].

Mathematical modeling has emerged as a critical tool for malaria intervention planning, enabling systematic evaluation of control strategies under diverse epidemiological scenarios [5, 6]. However, existing optimization approaches typically employ classical optimal control methods (e.g., Pontryagin’s Maximum Principle), which face computational limitations when applied to high-dimensional, multi-constraint problems with nonlinear dynamics [7]. Prior work on *P. vivax* has also incorporated relapse dynamics [2, 8] and climate-driven vector traits [9, 10], but few studies combine these elements in a policy-driven optimization framework.

In Korea, several modeling studies have addressed seasonal relapse distributions, rapid diagnostic testing, and regional surveillance systems [11, 12]. These highlight the operational challenges of *P. vivax* elimination but do not perform structural identifiability analysis or optimize monthly intervention timing under supply constraints. Meanwhile, recent tafenoquine cost-effectiveness studies [13, 14] demonstrate favorable relapse prevention compared to primaquine but do not solve a high-dimensional intervention scheduling problem that integrates stockpile allocation and seasonal risk.

Against this backdrop, our study advances the literature in several ways. First, we perform a structural identifiability (SI) analysis tailored to a *P. vivax* relapse- and climate-aware model, mapping observables to identifiable and fixed parameter sets. While SI has gained traction in infectious disease modeling [15, 16], it remains rare in applied *P. vivax* transmission studies. Second, we address a 48-dimensional, month-by-month intervention scheduling problem using the Improved Multi-Operator Differential Evolution (IMODE) algorithm, which can robustly handle discrete, nonconvex constraints that classical control methods cannot. Third, we explicitly model stockpile and acceptability constraints for tafenoquine deployment, a novel feature that links supply-chain realities to seasonal optimization. Finally, we integrate relapse-focused cost-benefit metrics (IBCR, NPV) under Seoul’s elimination program, providing climate-adaptive recommendations for tafenoquine deployment and non-pharmaceutical interventions.

This study addresses these challenges by developing an integrated mathematical approach that combines *P. vivax* transmission modeling with advanced metaheuristic optimization. We employ the Improved Multi-Operator Differential Evolution (IMODE) algorithm to solve high-dimensional intervention optimization problems under climate change scenarios. The approach incorporates SI analysis to ensure reliable parameter estimation and evaluates both pharmaceutical and non-pharmaceutical interventions across multiple climate scenarios.

Our specific objectives are: (I) develop and validate a *P. vivax* transmission model incorporating hypnozoite dynamics and climate-dependent vector parameters; (II) optimize intervention strategies using IMODE algorithm under realistic resource constraints; (III) evaluate economic cost-benefit of tafenoquine deployment strategies; and (IV) generate climate-adaptive intervention recommendations for Seoul metropolitan area. This case study demonstrates the methodology’s applicability for evidence-based malaria control planning.

## 2. Mathematical model and parameter analysis

The mathematical model incorporates the distinctive features of *P. vivax* transmission, including hypnozoite formation and temperature-dependent vector dynamics [17, 18]. Environmental factors, particularly temperature and precipitation, critically influence mosquito population dynamics [19, 20].

### 2.1. Mathematical model for malaria transmission

We developed a compartment model for the transmission dynamics of *Plasmodium vivax* malaria by distinguishing between human and mosquito populations. The human population is divided into susceptible (*S*_*H*_ ), exposed with short- and long-term latency (*E*_*HS*_ and *E*_*HL*_), infectious (*I*_*H*_ ), dormant liver-stage carriers (*D*_*H*_ ), relapse latency (*L*_*HS*_ and *L*_*HL*_), treated (*T*_*T*_ and *T*_*P*_ ), and recovered (*R*) hosts in the gray area. The mosquito population includes eggs in the aquatic stage (*A*), susceptible adults (*S*_*V*_ ), exposed (*E*_*V*_ ), and infectious (*I*_*V*_ ) mosquitoes in the purple area in Figure 1.

**Figure 1:**
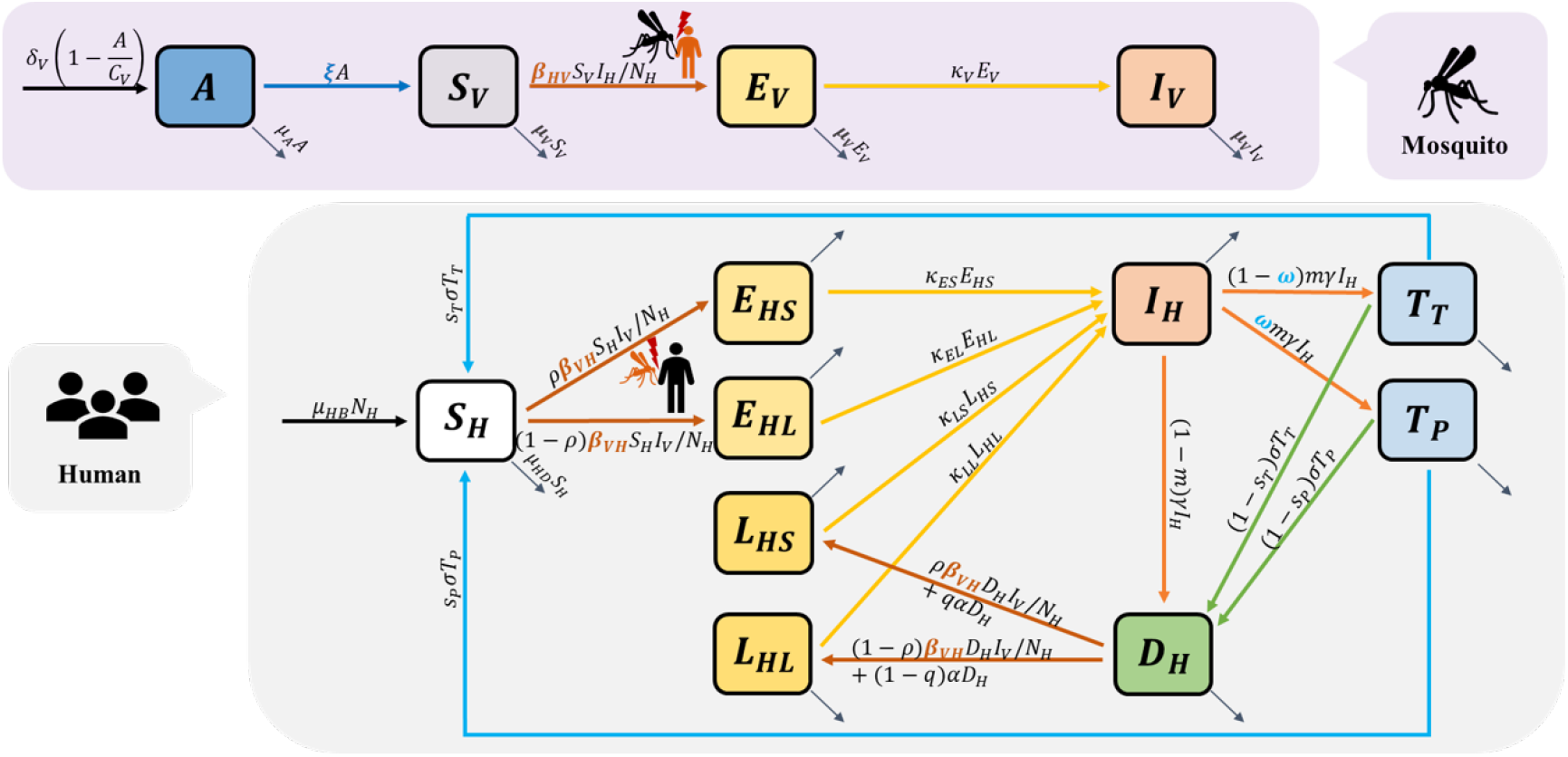
Compartmental structure of the Plasmodium vivax malaria transmission model. The model couples human and mosquito populations with temperature- and precipitationdependent parameters. Human compartments include susceptible (*S*_*H*_ ), exposed with short and long latency (*E*_*HS*_, *E*_*HL*_), infectious (*I*_*H*_ ), dormant hypnozoites (*D*_*H*_ ), latent relapse stages (*L*_*HS*_, *L*_*HL*_), treated with tafenoquine (*T*_*T*_ ) or primaquine (*T*_*P*_ ), and recovered (*R*). Mosquito compartments include aquatic stage (*A*), susceptible adults (*S*_*V*_ ), exposed (*E*_*V*_ ), and infectious (*I*_*V*_ ). Environmental forcing shapes mosquito maturation, mortality, and biting rates. Model observables include: (i) confirmed human cases (from latent to infectious transitions), (ii) treated human cases, (iii) infectious mosquitoes, (iv) susceptible mosquitoes, (v) total human population, and (vi) total mosquito population. These observables form the basis for SI analysis, while only (i) confirmed human cases are used for model calibration and parameter estimation.

The transmission cycle begins with mosquito eggs in aquatic habitats developing into adult mosquitoes at temperature-dependent maturation rates, with active development occurring between 20-30°C and precipitation levels of 10-40mm. Adult mosquitoes become infected after feeding on infectious humans (*I*_*H*_ ) or dormant carriers (*D*_*H*_ ), then undergo an extrinsic incubation period (1*/κ*_*V*_ ) before becoming capable of transmission. Infectious mosquitoes can then transmit parasites to susceptible humans, who progress through either short-term (1*/κ*_*ES*_) or long-term (1*/κ*_*EL*_) latency periods before becoming infectious. A characteristic feature of *P. vivax* is the formation of dormant liver-stage parasites (hypnozoites) following treatment. Infectious individuals receive antimalarial treatment, with a probability *ω* of receiving primaquine and probability (1 − *ω*) of receiving tafenoquine. Treatment efficacies for eliminating hypnozoites (*s*_*T*_ and *s*_*P*_ ) determine whether individuals recover completely or enter the dormant state (*D*_*H*_ ). Dormant infections can reactivate through relapse mechanisms, following short-term (1*/κ*_*LS*_) or long-term (1*/κ*_*LL*_) latency periods before returning to the infectious state. Table 2 presents the temperature and precipitation dependent parameters for mosquito life cycle processes. Our model structure is illustrated in Figure 1, with biological and epidemiological parameters listed in Table 1. The system dynamics are governed by the following ordinary differential equations:

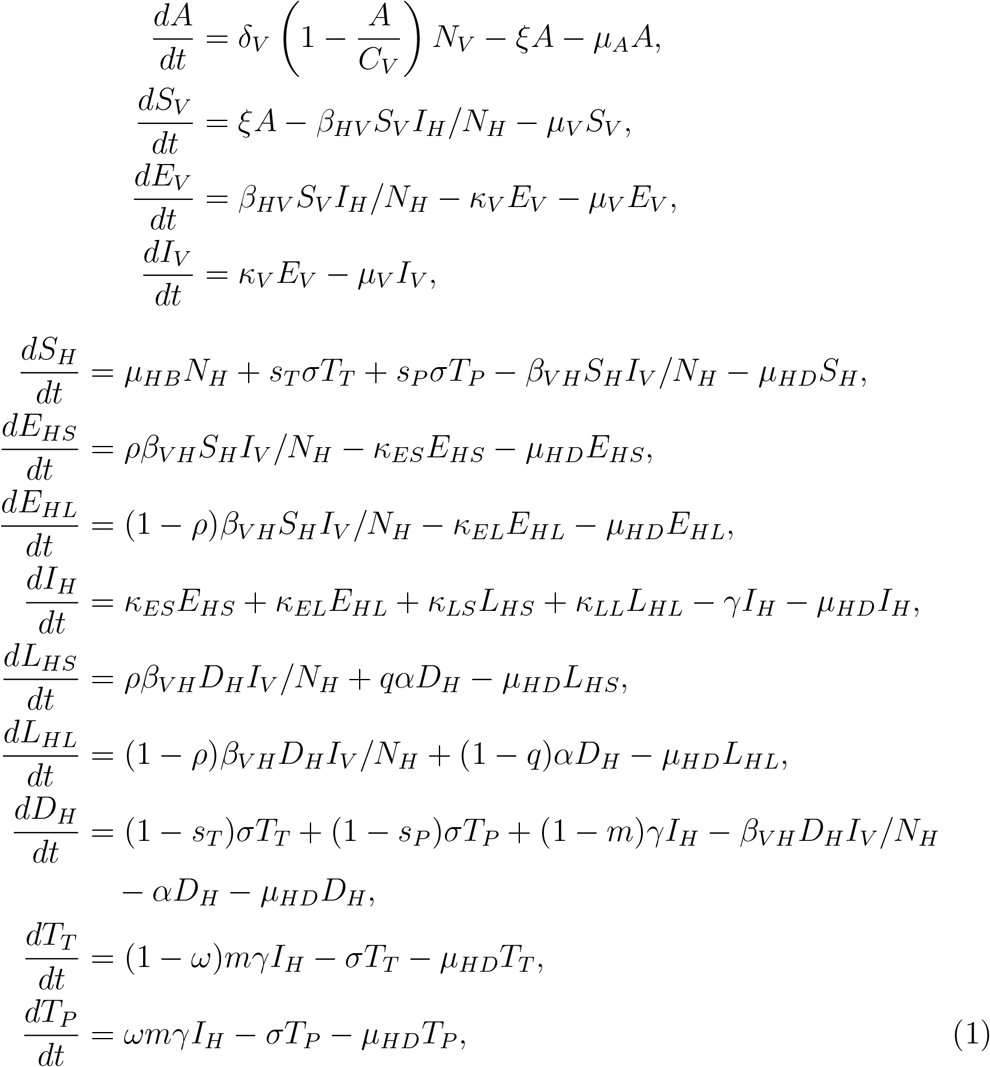

where *N*_*H*_ = *S*_*H*_ +*E*_*HS*_ +*E*_*HL*_ +*I*_*H*_ +*L*_*HS*_ +*L*_*HL*_ +*D*_*H*_ +*T*_*T*_ +*T*_*P*_ represents the total human population and *N*_*V*_ = *S*_*V*_ + *E*_*V*_ + *I*_*V*_ represents the total adult mosquito population.

**Table 1:** Time-independent parameters in the *P. vivax* malaria model, including temperature-dependent transmission rates (*β*_*HV*_, *β*_*V H*_ ), relapse and latency parameters (*ρ*, 1*/α*), and treatment efficacy terms (*m, ω, s*_*T*_, *s*_*P*_ ).

| Symbol | Description | Value | Reference |
| --- | --- | --- | --- |
| $\beta_{HV}$ | Transmission rate (human to mosquito) | $b_V b(T)$ | [21, 22, 23] |
| $\beta_{VH}$ | Transmission rate (mosquito to human) | $b_H b(T)$ | [21, 22, 23] |
| $b_V$ | Probability that a susceptible mosquito becomes infected after biting an infected human | 0.0452 | Estimated |
| $b_H$ | Probability that a susceptible human becomes infected after being bitten by an infected mosquito | 0.4996 | Estimated |
| $C_V$ | Carrying capacity of adult mosquitoes | $S_H(0) \times 5$ | [24] |
| $1/\kappa_V$ | Average extrinsic latent period in mosquitoes | 10 days | [25] |
| $\mu_{HB}$ | Birth rate of humans | $1.5068 \times 10^{-6}$ | [26] |
| $\mu_{HD}$ | Mortality rate of humans | $1.5068 \times 10^{-6}$ | [26] |
| $\rho$ | Proportion with short-term latency period after infection | 0.641 | [27] |
| $1/\kappa_{ES}$ | Short-term latent period in humans | 25.5 days | [27] |
| $1/\kappa_{EL}$ | Long-term latent period in humans | 329.4 days | [27] |
| $1/\gamma$ | Average infectious period in humans | 14 days | [28] |
| $1/\kappa_{LS}$ | Short-term latent period of relapse | 14 days | [29, 30] |
| $1/\kappa_{LL}$ | Long-term latent period of relapse | 274 days | [29, 30] |
| $q$ | Proportion of cases with short-term latent period | 0.75 | [31] |
| $1/\alpha$ | Average relapse period in humans | 150 days | [32] |
| $m$ | Probability of receiving antimalarial treatment | 0.87 | [31] |
| $\omega$ | Probability of receiving primaquine | 1 | [31] |
| $s_T$ | Probability of liver-stage clearance by tafenoquine | 0.919 | [33] |
| $s_P$ | Probability of liver-stage clearance by primaquine | 0.773 | [33] |
| $1/\sigma$ | Average duration of drug effectiveness | 35 days | [34] |

**Table 2:** Time-dependent parameters for mosquito population dynamics. All temperaturedependent functions are defined for realistic temperature ranges (T in °C) and precipitation values (R in mm). Parameters control egg-laying, development, survival, and feeding behavior of Anopheles vectors under varying climatic conditions.

| Symbol | Description | Value | Reference |
| --- | --- | --- | --- |
| $\delta_V(T)$ | Egg-laying rate of female mosquitoes | $\max(-0.153T^2 + 8.61T - 97.7, 0)$ | [24] |
| $\xi(T, R)$ | Maturation rate | $\frac{e(T)p_E(R)p_L(R)p_P(R)}{\tau_{EA}(T)}, 16.5 \leq T \leq 35.6$<br>$e(T) = \frac{1}{2\pi} \exp\left(-\frac{(T-\mu)^2}{2\sigma^2}\right), \mu = 8, \sigma = 0.5$<br>$p_E(R) = -0.153T^2 + 8.61T - 97.7$<br>$p_L(R) = \frac{1}{1+e^{-0.00564(T-16.5)}}$<br>$p_P(R) = \min(1, \max(0, 0.84 + 0.067T))$<br>$\tau_{EA}(T) = \frac{298.5+1}{0.00031T^2-0.0154T+0.04}$ | [21, 24, 36] |
| $\mu_A(T)$ | Mortality rate of mosquito eggs | $\min\left(0.002 \exp\left(\left(\frac{T-23.7}{5.95}\right)^2\right), 1\right)$ | [35, 24] |
| $\mu_V(T)$ | Mortality rate of adult mosquitoes | $\max(-\ln(-0.0008287T^2 + 0.0367T + 0.522), 0)$ | [35, 24] |
| $b(T)$ | Biting rate | $0.0002037(T - 11.7)\sqrt{42.3 - T}$ | [21, 22, 23] |

### 2.2. Structural identifiability

SI analysis determines whether model parameters can be uniquely estimated from observed data [38, 39]. We examined two scenarios: “Known initial conditions” where all compartment initial values are assumed known, and “Unknown initial conditions” where they are treated as unknown additional parameters. In our parameter estimation (Section 2.3), all initial conditions were fixed using surveillance data and ecological baseline values, corresponding to the “Known initial conditions” scenario.

In our mathematical model for malaria transmission, the observable outputs were defined as

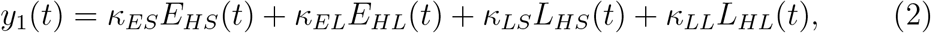

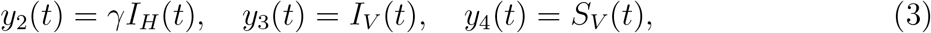

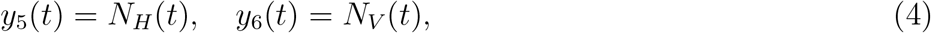

where *y*_1_(*t*) represents the number of confirmed cases, *y*_2_(*t*) represents the number of treatment cases, *y*_3_(*t*) represents the number of infected mosquitoes, *y*_4_(*t*) represents the number of susceptible mosquitoes, and *y*_5_(*t*), *y*_6_(*t*) represent total human and mosquito populations, respectively. Among these theoretical observables, *y*_1_(*t*) (confirmed cases) is directly measurable through confirmed data. Mosquito population data (*y*_3_(*t*), *y*_4_(*t*)) can be estimated through vector surveillance and trapping programs, though with greater uncertainty than human case data.

The SI analysis employs differential algebra methods by repeatedly differentiating observation functions *y* = *h*(*x*(*t*), *θ*) with respect to time, substituting state dynamics *x*(*t*)′ = *f* (*x*(*t*), *θ*), and eliminating unobserved states.

This process yields input-output polynomial relations *P* (*y, y*′, *y*′′, … ; *θ*) = 0 where the polynomial coefficients depend on the model parameters. Parameters can be classified into three categories: globally identifiable if they can be uniquely determined from observations, locally identifiable if only a finite number of values are consistent with the data, and non-identifiable if infinitely many values produce identical outputs. As an example, the transmission parameter *β*_*HV*_ can be shown to be globally identifiable. From the observable infected mosquito dynamics *y*_3_(*t*) = *I*_*V*_ (*t*),

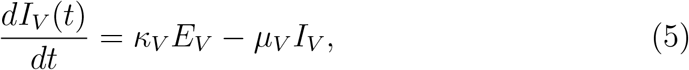

we can express the unobserved exposed mosquitoes as

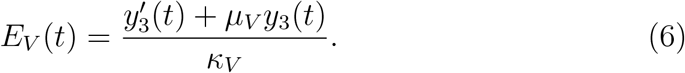

Substituting into the exposed mosquito equation

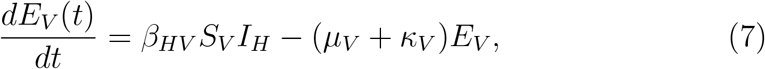

with *S*_*V*_ = *y*_4_, *I*_*H*_ = *y*_2_*/γ*, yields

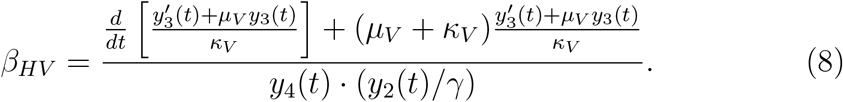

In contrast, the mosquito carrying capacity *C*_*V*_ appears in the mosquito population dynamics as:

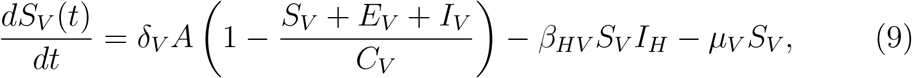

but since the aquatic stage *A* and exposed mosquitoes *E*_*V*_ are unobserved, *C*_*V*_ cannot be separated from the total mosquito population ratio (*S*_*V*_ +*E*_*V*_ + *I*_*V*_ )*/C*_*V*_ . This structural coupling makes *C*_*V*_ non-identifiable, justifying its fixation to literature values.

The SI analysis was performed using the Julia package `StructuralIdentfiability.jl` [37]. Results are summarized in Table 3, showing that key epidemiological parameters (*β*_*HV*_, *β*_*V H*_, *γ, m, ω, s*_*P*_, *s*_*T*_, *σ, µ*_*HB*_, *µ*_*HD*_) are structurally identifiable, while ecological mosquito parameters (*C*_*V*_, *δ*_*V*_, *µ*_*A*_, *ξ*) remain non-identifiable due to coupling with unobserved states. This identifiability analysis provides the theoretical foundation for our parameter estimation approach. We estimate globally identifiable transmission parameters (*β*_*HV*_, *β*_*V H*_ ) from data as described in Section 2.3, while fixing non-identifiable parameters to well-established values using literature values and conducting the sensitivity analysis. Section 2.4 subsequently quantifies the relative influence of all parameters on transmission dynamics to validate this estimation strategy.

**Table 3:** Structural identifiability of the model parameters under different assumptions about initial conditions for compartments. In the “Known initial conditions” scenario, all compartments except *A* and *E*_*V*_ are assumed to have known initial values.

| Parameter | Known initial conditions | Unknown initial conditions |
| --- | --- | --- |
| $\beta_{HV}, \beta_{VH}$ | Globally identifiable | Globally identifiable |
| $\kappa_V, \mu_{HB}, \mu_{HD}, \gamma, \alpha, m, \omega, s_P, s_T, \sigma$ | Globally identifiable | Globally identifiable |
| $\kappa_{ES}, \kappa_{EL}, \kappa_{LS}, \kappa_{LL}, q, \rho$ | Globally identifiable | Locally identifiable |
| $C_V, \mu_A, \mu_V$ | Globally identifiable | Non-identifiable |
| $\delta_V, \xi$ | Non-identifiable | Non-identifiable |

### 2.3. Model calibration

Parameters directly informed by Seoul-specific data include relapse periodicity and treatment allocation between primaquine and tafenoquine, reflecting the national malaria elimination program context. Assumptions on drug efficacy were aligned with local programmatic use but informed primarily by international clinical studies. In contrast, entomological parameters (temperature- and precipitation-dependent maturation, adult mortality, and carrying capacity) were taken from global studies due to limited Korea-specific estimates. We tested the influence of these global estimates using global sensitivity analysis and PRCC (Partial Rank Correlation Coefficients), confirming that our main conclusions are robust to uncertainty in these values.

#### 2.3.1. Data sources

Our analysis draws on two primary data sources: (i) epidemiological surveillance data on *Plasmodium vivax* malaria in Seoul, Republic of Korea, and (ii) local climate data used to parameterize temperature- and precipitationdependent mosquito dynamics. Weekly confirmed malaria case counts were obtained from the Seoul Metropolitan Government surveillance system for the period May 1 to December 31, 2023. These data provide the observed incidence time series used for parameter estimation and model calibration. Treatment coverage rates with primaquine and tafenoquine, as well as relapse periodicity, were informed by national surveillance reports and the Korean Disease Control and Prevention Agency (KDCA) [40]. Climate variables were extracted from the Korea Climate Information Portal [41], including daily temperature and precipitation values for Seoul. These time series were incorporated into the model to drive the temperature- and rainfall-dependent mosquito maturation, mortality, and biting rate functions. Together, these epidemiological and climate data provide the empirical foundation for calibrating the transmission model, testing parameter identifiability, and evaluating the impact of intervention strategies in the context of Seoul’s malaria elimination program.

#### 2.3.2. Parameter estimation and parametric bootstraping

Based on the SI analysis, we estimated the globally identifiable parameters *β*_*HV*_ and *β*_*V H*_ using weekly *P. vivax* malaria confirmed data from Seoul, Korea from May 1 to December 31, 2023. The observed data consists of weekly confirmed cases reported through the Seoul Metropolitan Government surveillance system, with actual temperature and precipitation data incorporated into the temperature-dependent mosquito parameters.

Let *θ* = [*β*_*HV*_, *β*_*V H*_ ]^*T*^ ∈ Ω = [0, 1] × [0, 1] denote the parameter vector. Define the forward operator *F* : Ω → ℝ^35^ that maps parameters to weekly model predictions via the *P. vivax* transmission model:

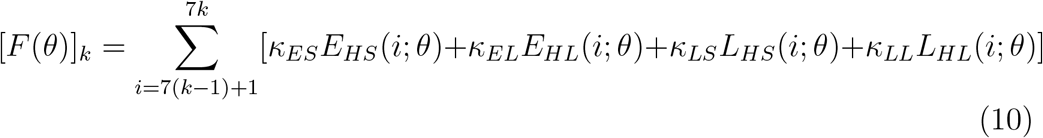

where *E*_*HS*_, *E*_*HL*_, *L*_*HS*_, *L*_*HL*_ represent the exposed human compartments from the ODE system in (1), and *k* = 1, 2, …, 35 denotes weekly time points. The parameter estimation is formulated as the least-squares problem:

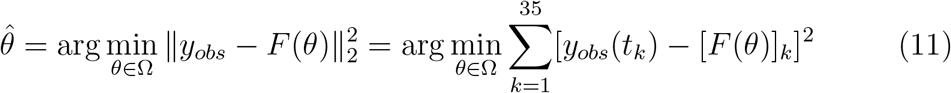

The parametric bootstrap procedure for uncertainty quantification follows [42]: Given the estimated parameters 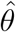, we generate *B* = 1000 synthetic datasets 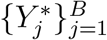 where 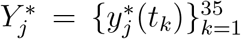 with 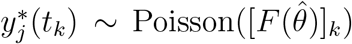. This approach assumes that observed weekly case counts follow Poisson distributions around the model predictions, which is appropriate for epidemiological count data with relatively low incidence rates. For each synthetic dataset 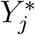, we solve the parameter estimation problem:

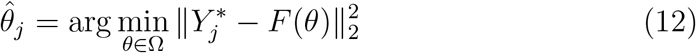

using the same least-squares optimization procedure applied to the original data. The resulting bootstrap parameter ensemble 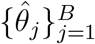 captures the sampling distribution of the parameter estimates under the assumed noise model. The bootstrap predictions 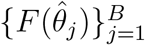 provide confidence intervals for weekly case projections,, with detailed parameter distributions presented in Appendix B.

Figure 2 shows the model fitting results with observed weekly confirmed data and bootstrap distribution at the final time point. The model demonstrates good agreement with observed data (*R*^2^ = 0.75), and the bootstrap distribution provides 95% confidence intervals of [89, 124] cases.

**Figure 2:**
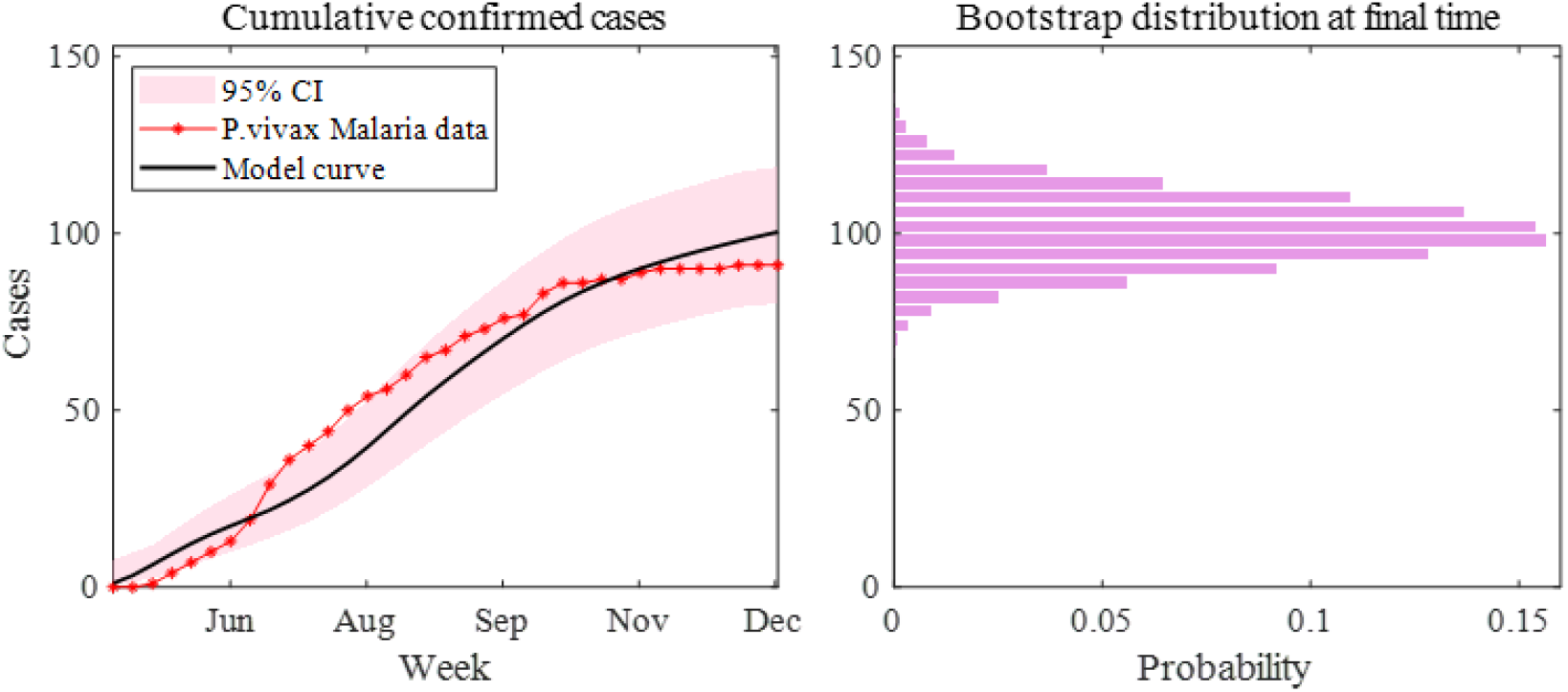
Model fitting to Seoul P. vivax confirmed data and bootstrap uncertainty analysis. Left panel shows observed weekly cases with model predictions. Right panel displays bootstrap distribution of predicted cases at final time point.

### 2.4. Sensitivity analysis

Having calibrated identifiable transmission parameters in Section 2.3, we assess the relative importance of all model parameters on transmission dynamics. This analysis serves two purposes. First, it validates that estimated parameters exert dominant influence on model predictions. Second, it quantifies the potential impact of uncertainty in fixed non-identifiable parameters on model behavior.

We employ two complementary approaches tailored to parameter characteristics. For time-dependent climate-modulated parameters (*δ*_*V*_ (*T* ), *µ*_*V*_ (*T* ), *b*(*T* ), *µ*_*A*_(*T* ), *ξ*(*T, R*)), we use PRCC analysis to assess their influence under realistic climate variation while accounting for temporal correlations. For time-invariant parameters, including both estimated (*β*_*HV*_, *β*_*V H*_ ) and fixed (*C*_*V*_, etc.) values, we use Sobol indices to decompose output variance into individual contributions and interactions. This dual approach provides comprehensive insight into both climate-driven seasonal dynamics and intrinsic transmission mechanisms.

#### 2.4.1. Partial Rank Correlation Coefficient (PRCC)

Climate-driven parameters exhibit strong seasonal variation directly modulated by temperature and precipitation. These time-dependent inputs require sensitivity analysis methods that can handle temporally correlated parameter trajectories while quantifying monotonic relationships with model outputs [43]. We employed PRCC analysis with Latin Hypercube Sampling (LHS) to efficiently explore the parameter space and assess the relative influence of each climate-modulated parameter on transmission dynamics.

Latin Hypercube Sampling [44] generates parameter sets by partitioning each parameter’s range into equal probability intervals and sampling once from each interval, ensuring comprehensive coverage of the parameter space with minimal sample size. For each time-dependent parameter *p*_*i*_(*t*), we generated *N* = 10000 samples from uniform distributions centered on baseline values with *±*20% variation ranges. The PRCC for parameter *p*_*i*_ measures the Spearman rank correlation between the model output *Y* and *p*_*i*_ after accounting for the linear effects of all other parameters **p**_∼*i*_:

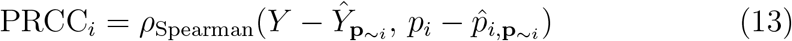

where 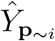 and 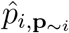 represent fitted values from linear regressions. PRCC values range from ሢ1 to +1, with magnitude indicating strength of monotonic association and sign indicating direction of influence. Statistical significance was assessed using bootstrap resampling (*n* = 10000 iterations).

Figure 3 displays climate-driven parameter trajectories (black lines, left axis) and corresponding PRCC values (shaded regions, right axis) across Seoul’s 2023 transmission season. The top-left panel shows temperature (−5 to 30°C) and precipitation (0-100 mm) inputs driving the parameter functions defined in Table 2. Parameter fluctuations exhibited distinct seasonal patterns. The egg-laying rate *δ*_*V*_ (*T* ) varied from 5 to 22 eggs/female/day with summer peaks. Adult mortality *µ*_*V*_ (*T* ) remained below 0.2 day^−1^ except during late-summer heat extremes, reaching up to 1.0 day^−1^. The maturation rate *ξ*(*T, R*) showed sporadic precipitation-driven peaks up to 3.0. The biting rate *b*(*T* ) ranged from near-zero below 15°C to 0.4 day^−1^ in midsummer. Egg mortality *µ*_*A*_(*T* ) increased from near-zero to 0.9 day^−1^ with rising temperatures.

**Figure 3:**
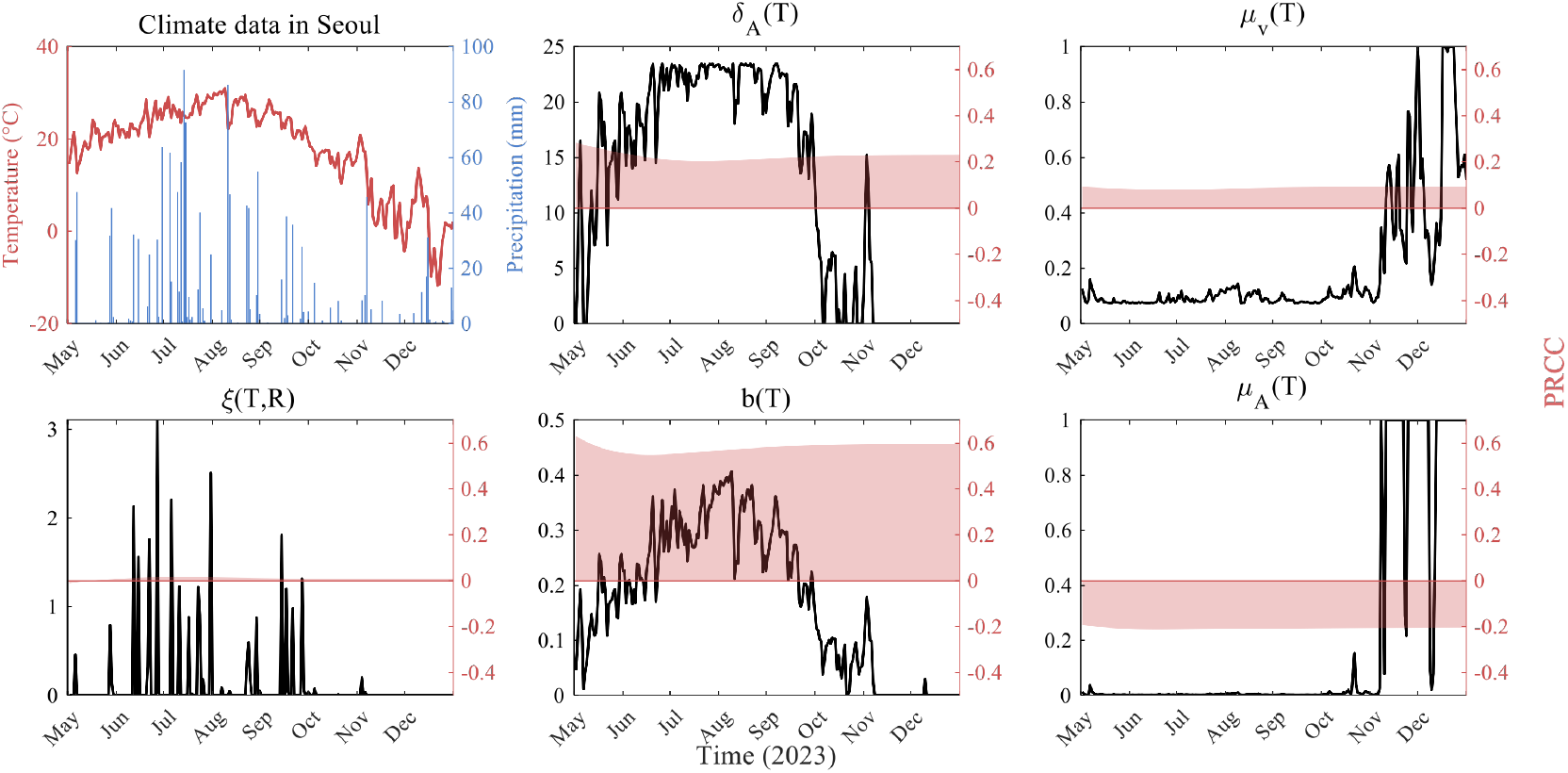
Time-dependent parameters across the 2023 Seoul transmission season. Panels show (left to right, top to bottom): temperature and precipitation inputs, egg-laying rate *δ*_*V*_ (*T* ), adult mortality *µ*_*V*_ (*T* ), maturation rate *ε*(*T, R*), biting rate *b*(*T* ), and egg mortality *µ*_*A*_(*T* ).

PRCC analysis revealed that *b*(*T* ) exerted dominant influence (PRCC = 0.4-0.6) throughout the transmission season, while all other parameters showed minimal sensitivity (|*PRCC*| *<* 0.2). The negative PRCC for *µ*_*A*_(*T* ) (approximately -0.2) indicates modest suppressive effects during peak season. These results demonstrate that adult mosquito biting behavior controls transmission dynamics under Seoul’s climate conditions. In contrast, aquatic-stage parameters and temperature-dependent mortality contribute negligibly to infection outcomes despite their substantial seasonal fluctuations.

#### 2.4.2. Sobol indices

Sobol sensitivity analysis decomposes the output variance into contributions from individual parameters (first-order indices) and parameter interactions (higher-order indices) [45]. We analyzed time-invariant parameters (*b*_*V*_, *b*_*H*_, *κ*_*V*_, *C*_*V*_, *γ*) using *N* = 10,000 samples (Saltelli scheme, *±*5–20% variation), with cumulative infections as output. The first-order Sobol index *S*_*i*_ quantifies the main effect of parameter *p*_*i*_ on model output variance:

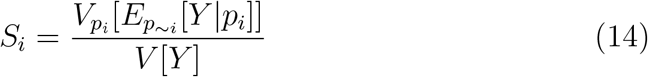

where 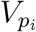 denotes variance with respect to parameter *p*_*i*_, 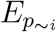 represents expectation over all parameters except *p*_*i*_, and *Y* is the model output. The total-order index captures both main effects and all interaction terms involving parameter *p*_*i*_:

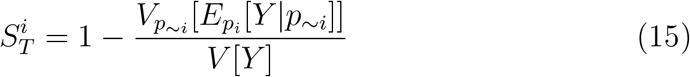

Figure 4 shows Sobol indices for cumulative infectious cases across the 2023 transmission season. The human infection probability *b*_*H*_ exhibits the highest sensitivity throughout the period, with values ranging from 0.8 in early summer (June-July) to 0.3-0.4 in late autumn (November-December), While remaining the most influential parameter throughout the season, its importance decreases substantially as temperatures decline and mosquito activity diminishes. The mosquito infection probability *b*_*V*_ (green) shows moderate and relatively stable sensitivity (0.1-0.2) with slight increases during peak transmission months. In contrast, the mosquito carrying capacity *C*_*V*_ (purple) and recovery rate *γ* (yellow) demonstrate consistently low sensitivity (*<* 0.1) across all time periods, indicating their limited influence on cumulative case predictions. The near-identical first-order (left) and total-order (right) indices indicate negligible parameter interactions, with individual parameter effects (main effects) accounting for over 95% of total sensitivity. This additive structure simplifies uncertainty quantification and parameter prioritization for malaria intervention strategies.

**Figure 4:**
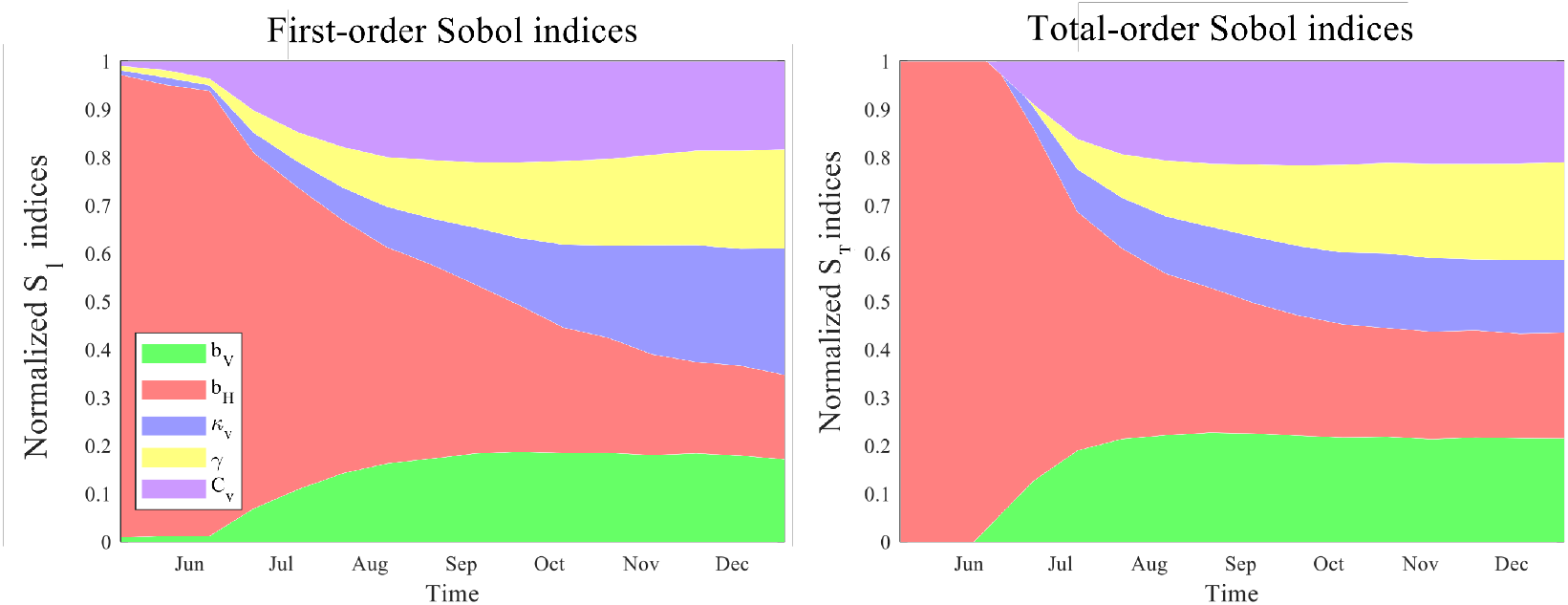
First-order (*S*_1_) and total-order (*S*_*T*_ ) Sobol indices for transmission parameters *b*_*V*_, *b*_*H*_, extrinsic incubation period *κ*_*V*_, mosquito carrying capacity *C*_*V*_, and human recovery rate *γ*.

## 3. Formulation of the optimization problem

### 3.1. Intervention strategies and climate change scenarios

Climate variability directly influences mosquito dynamics through temperaturedependent parameters, including the egg-laying rate *δ*_*V*_ (*T* ), maturation rate *ξ*(*T* ), adult mortality rate *µ*_*V*_ (*T* ), and biting rate *b*(*T* ). Based on climate data for Seoul, we consider three climate scenarios (S1-S3) representing different temperature and precipitation patterns in Figure 5.

**Figure 5:**
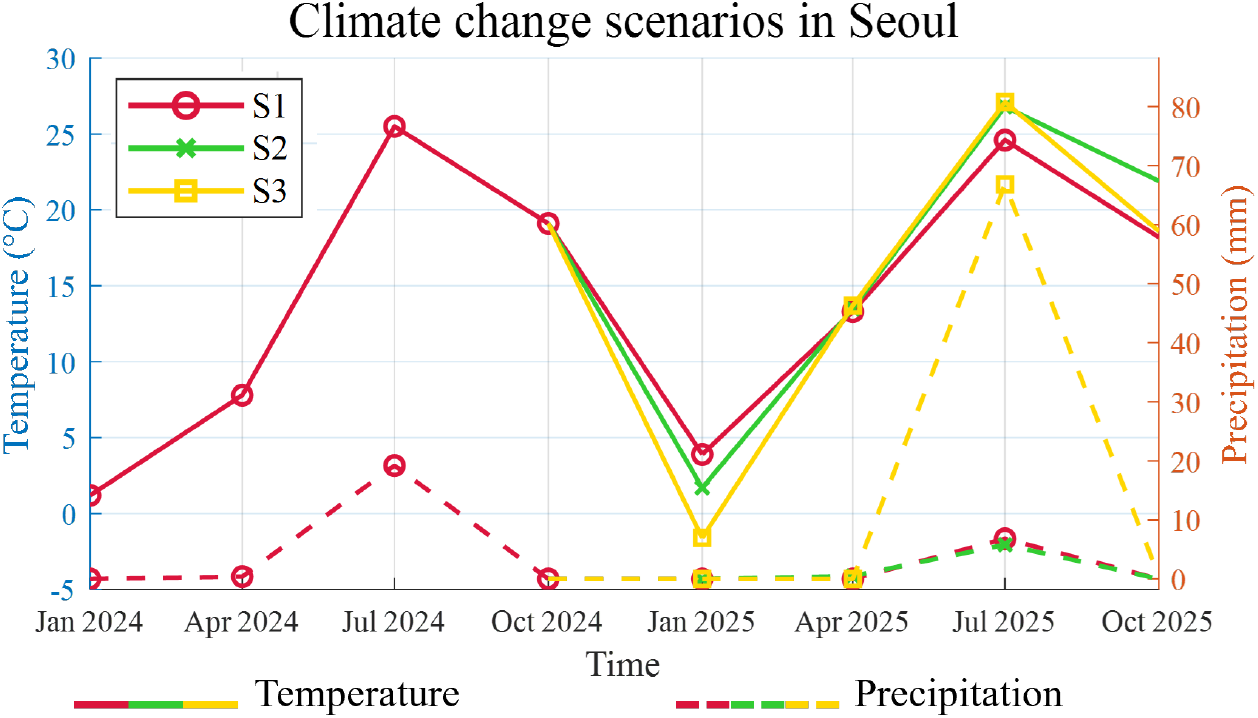
Climate scenarios for Seoul showing temperature and precipitation patterns at 3-month intervals.

We examine four types of malaria intervention strategies that can be optimized to minimize disease transmission:

- Human-mosquito contact reduction modifies the transmission rates *β*_*V H*_, *β*_*HV*_ as (1 − *c*_*β*_)*β*_*V H*_ and (1 − *c*_*β*_)*β*_*HV*_, where *c*_*β*_ represents the intensity of protective measures such as repellents, window screens, and bed nets.
- Larval control reduces the maturation rate of aquatic stages, expressed as (1 − *c*_*ξ*_)*ξ*, with *c*_*ξ*_ accounting for environmental management and larvicide application targeting mosquito breeding sites.
- Adult mosquito control increases the mortality rate of adult mosquitoes, modeled as 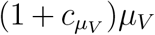, where 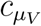 reflects interventions such as insecticide spraying and mosquito traps.
- Pharmaceutical intervention modifies the treatment regimen by substituting primaquine with tafenoquine. The probability of receiving primaquine *ω* is adjusted according to the control intensity *c*_*ω*_, resulting in a tafenoquine proportion of 1 − *ω*(1 − *c*_*ω*_). When *c*_*ω*_ = 1, primaquine is fully substituted by tafenoquine during the intervention period.

For the primary analysis, we focus on climate scenario S1 and evaluate all four intervention strategies. Under this scenario, the pharmaceutical intervention strategy is examined in detail by varying the maximum proportion of tafenoquine use across five levels (0, 0.25, 0.5, 0.75, and 1). Additional analyses of non-pharmaceutical interventions across climate scenarios S2 and S3 are given in Appendix C.

### 3.2. Objective function and constraints

We formulate the malaria intervention optimization as a constrained optimal control problem to minimize disease transmission while respecting practical implementation constraints.

The optimal control problem is to minimize the cumulative number of infectious humans:

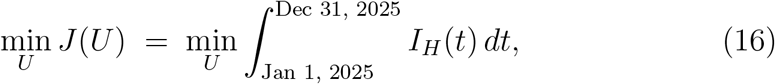

where *I*_*H*_ (*t*) denotes the number of infectious individuals at time *t*, and the control vector is

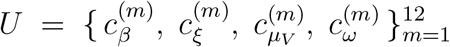

representing monthly control levels (*m* = 1, …, 12 for January–December 2025) for human-mosquito contact reduction, larval control, adult mosquito control, and pharmaceutical intervention, respectively. Each control is implemented as a piecewise-constant function over its corresponding month:

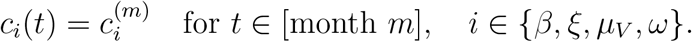

The admissible controls are subject to the bounds

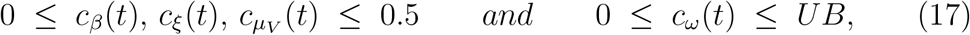

where *UB* ∈ {0, 0.25, 0.5, 0.75, 1} specifies the maximum monthly intensity of tafenoquine usage (Note that *UB*=0 implying no tafenoquine and *UB*=1 implying full substitution of primaquine).

Feasibility is enforced through two types of constraints. First, an acceptability constraint limits the annual cumulative intensity of each control:

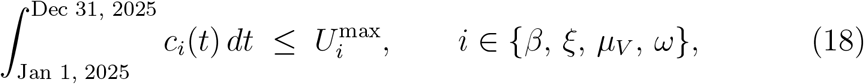

where 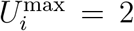 for all intervention types. Second, a stockpile constraint ensures that tafenoquine usage does not exceed available supply:

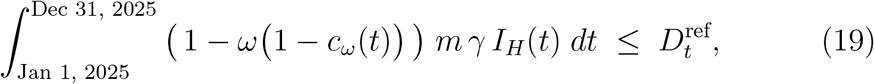

where 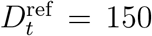 doses represents the available tafenoquine stockpile, assuming 20% substitution of primaquine treatment among the projected *P. vivax* patient population under primaquine-only treatment stratedy (baseline scenario, SA).

## 4. Solution method: IMODE algorithm

To solve the constrained optimal control problem defined in Section 3, we employ the Improved Multi-Operator Differential Evolution (IMODE) algorithm [46], implemented in MATLAB R2024b. IMODE was selected over classical optimal control methods due to the problem’s 48 discrete monthly decisions, nonconvex feasible regions, and non-differentiable penalty functions. Unlike classical optimal control methods that require analytical tractability, IMODE’s multi-operator framework with adaptive parameter control effectively handles the high-dimensional constraint space (48 decision variables) and nonconvex feasible region characteristic of our malaria intervention problem. The algorithm’s proven robustness on challenging optimization benchmarks, combined with its constraint-handling capabilities through penalty methods, makes it well-suited for this epidemiological control application. The malaria intervention problem exhibits these characteristics due to the complex nonlinear dynamics of the underlying epidemiological model and the multiple competing objectives implicit in the constraint structure.

The malaria transmission dynamics incorporate the control variables through the following transformations:

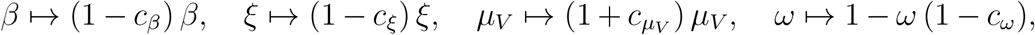

where *β* represents the transmission rate (*β*_*V H*_, *β*_*HV*_ ), *ξ* the mosquito maturation rate, and *µ*_*V*_ the adult mosquito mortality rate.

Constraint handling is implemented through a penalty-augmented fitness function:

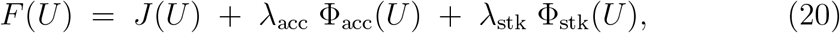

where 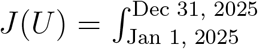 *I*_*H*_ (*t*) *dt* is the primary objective, and the penalty terms are defined as:

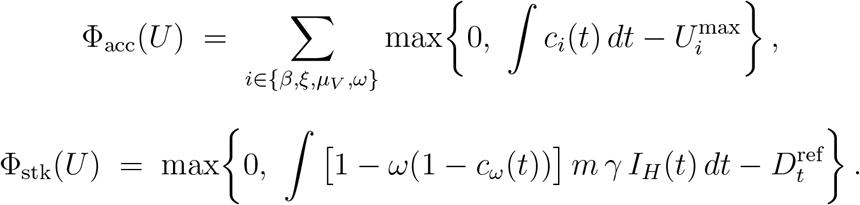

In our numerical experiments, we use large penalty weights (*λ*_acc_ = 10^5^ and *λ*_stk_ = 10^6^) to ensure that constraint violations are heavily penalized and feasible solutions are strongly preferred.

IMODE employs three mutation operators with adaptive parameter control and operator selection based on performance history. The algorithm evolves a population through mutation, crossover, and selection processes, with the procedure outlined in Algorithm 1.

## 5 Simulation results for optimal interventions

We applied the IMODE algorithm to determine optimal intervention strategies for *P. vivax* malaria control in Seoul. Figure 6 shows the optimal intervention profiles obtained from the IMODE optimization. The results reveal distinct temporal patterns with maximum intervention intensities not exceeding 0.5. Contact reduction measures (*C*_*β*_(*t*)) are optimally implemented from July to September, peaking at 0.5 intensity during July-August when transmission risk is highest. Larval growth suppression (*C*_*ξ*_(*t*)) follows a similar pattern from June to August, with optimal intensity around 0.45-0.5. Adult mosquito elimination 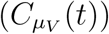 shows two distinct intervention periods: early spring (March-April) targeting diapausing mosquitoes, and mid-summer (July-September) for active population control. The early spring intervention provides a critical opportunity to reduce the founding population before the transmission season begins. The tafenoquine usage proportion (*C*_*ω*_(*t*)) demonstrates complex temporal patterns that vary significantly based on supply constraints. Under unrestricted availability (upper bound = 1.0), optimal deployment concentrates during peak transmission months (July-August), reaching maximum usage levels. When supply is constrained to 75% of demand, distribution extends from June to October with gradually declining allocation. Under a 25% supply limit, usage spreads across March to November, following seasonal transmission intensity patterns. It is important to highlight that the allocation strategies reflect the optimization algorithm’s response to both epidemiological factors and resource limitations. The optimal strategies align with the 2024 government malaria control plan [47], particularly the timing of surveillance and larval control activities (Figure 6, colored arrows).

**Figure 6:**
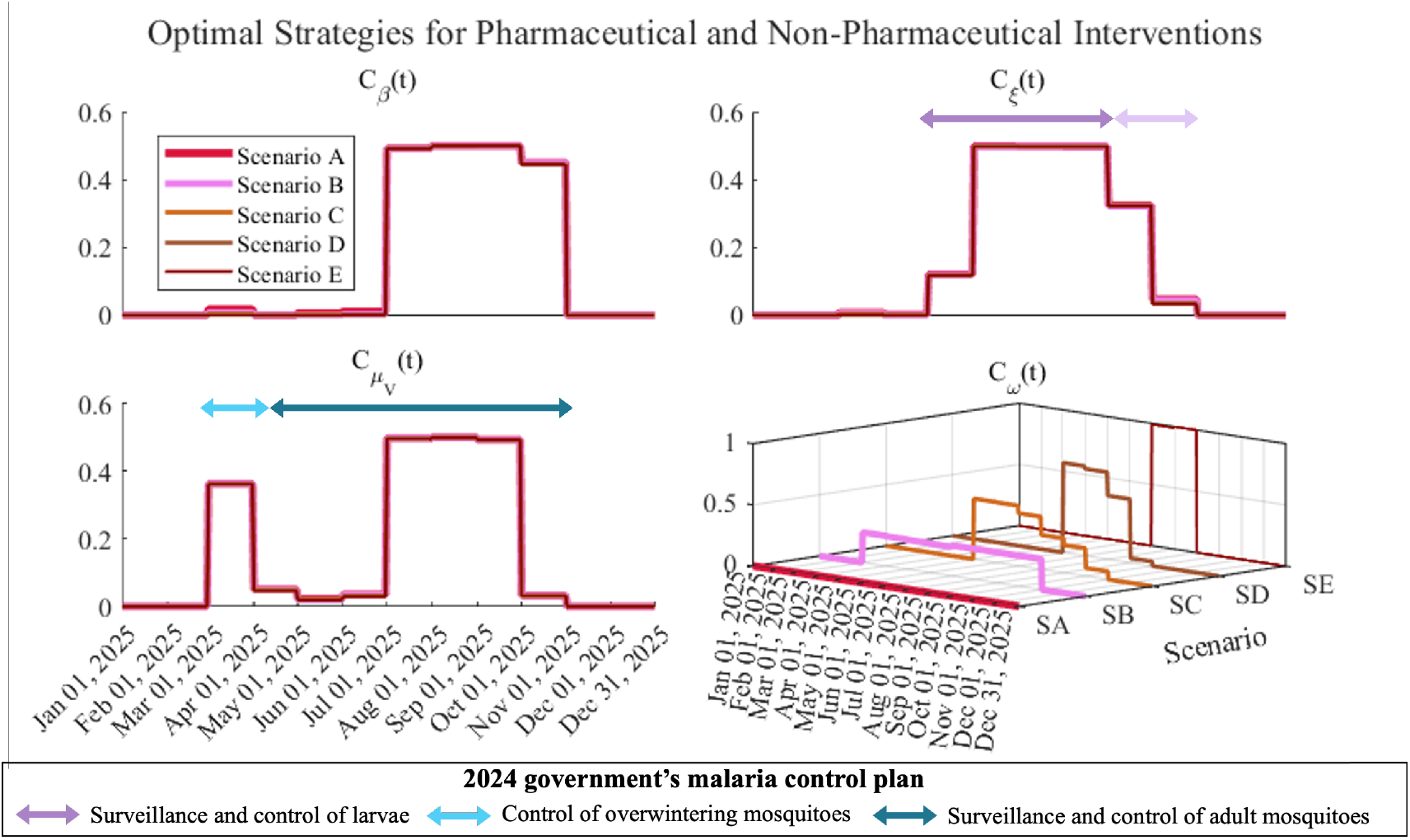
Optimal intervention strategies for pharmaceutical and non-pharmaceutical controls across different tafenoquine supply scenarios. Top panels show contact reduction (*C*_*β*_(*t*)) and larval control (*C*_*ξ*_(*t*)) strategies. Bottom left shows adult mosquito control 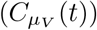 with early spring targeting of overwintering mosquitoes and summer population control. Bottom right displays the temporal evolution of tafenoquine usage proportion (*C*_*ω*_(*t*)) across scenarios SA-SE. Colored arrows(purple and blue) indicate alignment with Korea’s 2024 government malaria control plan for surveillance and vector management activities.

### Algorithm 1 IMODE algorithm for malaria intervention optimization

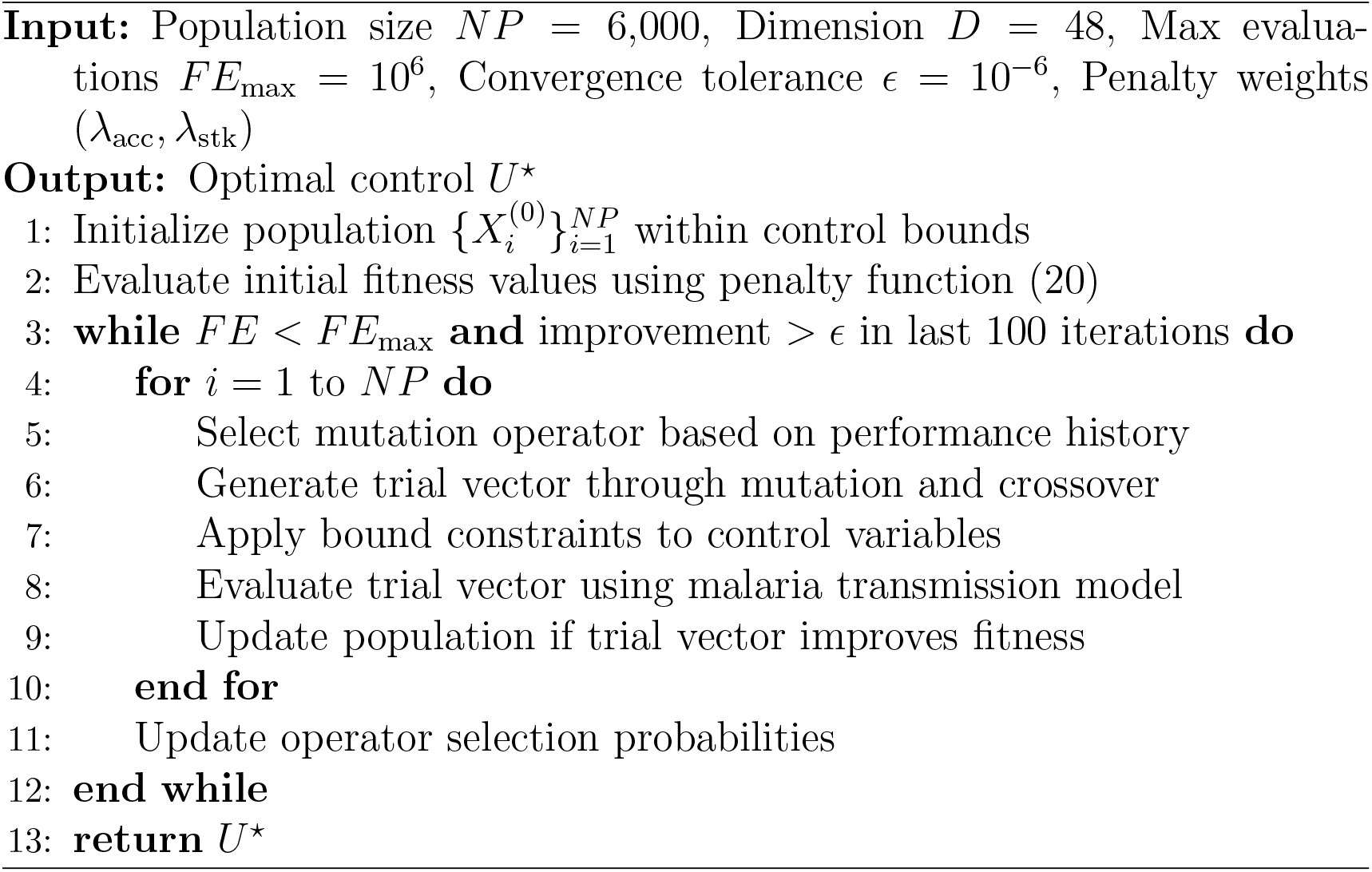

## 6. Cost–benefit evaluation

### 6.1. Numerical analysis across scenarios

Figure 7 presents the numerical results of five intervention scenarios (SASE) with varying tafenoquine coverage levels. Each scenario was evaluated using the IMODE optimization algorithm to determine optimal resource allocation under different supply constraints. The baseline scenario SA uses 100% primaquine treatment with optimized non-pharmaceutical interventions (contact reduction, larval control, adult mosquito control) determined by IMODE, resulting in approximately 850 total malaria cases, with 45 relapsed cases and zero prevented relapses. As tafenoquine upper bounds increase, total case numbers decrease progressively. Scenario SB with 25% coverage reduces cases to 780, while SC with 50% coverage achieves 760 cases. Scenario SD with 75% coverage further reduces cases to 750, and SE with 100% coverage reaches 740 cases. This represents an 18% reduction in total cases when comparing the unrestricted tafenoquine scenario to baseline.

**Figure 7:**
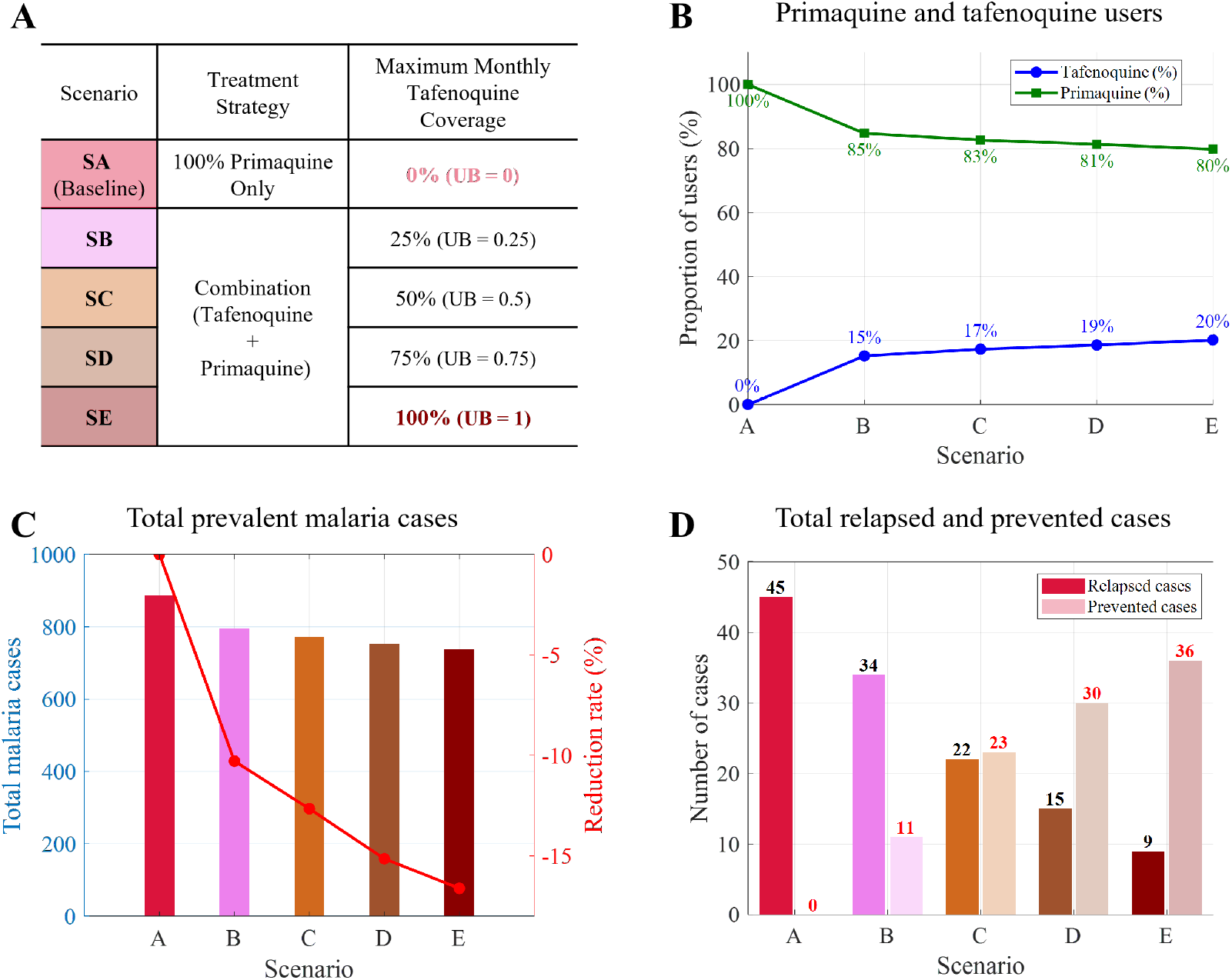
Comparative analysis of tafenoquine deployment scenarios showing epidemiological and operational outcomes. (A) Scenario definitions with treatment strategies and maximum monthly tafenoquine coverage limits (UB = upper bound). (B) Optimal drug allocation showing the proportion of tafenoquine users (blue line) and primaquine users (green line) resulting from IMODE optimization under different supply constraints. (C) Total prevalent malaria cases across scenarios with reduction rates relative to baseline (red line, right axis). (D) Relapse dynamics showing prevented cases (light bars) and remaining relapsed cases (dark bars) for each scenario.

The relapse prevention results demonstrate the most significant quantitative improvements. Scenario SE achieves 36 prevented cases with only 9 relapsed cases, representing an 80% reduction in relapse incidence compared to baseline. Intermediate scenarios show proportional benefits. SB prevents 11 cases with 34 relapses, SC prevents 23 cases with 22 relapses, and SD prevents 30 cases with 15 relapses. The results indicate that relapse prevention effectiveness increases nonlinearly with tafenoquine coverage.

### 6.2 Economic assessment of interventions

The economic evaluation quantifies the cost-effectiveness of different tafenoquine deployment strategies using standard pharmacoeconomic methods:

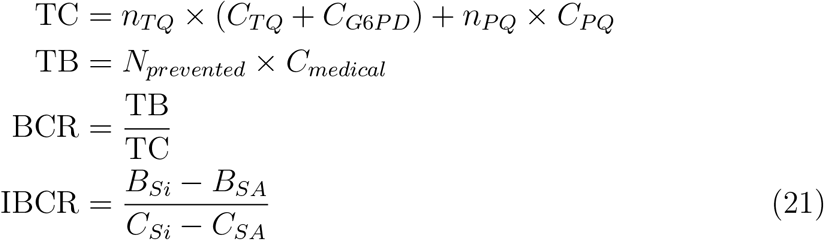

where *n*_*T Q*_ and *n*_*PQ*_ represent the number of tafenoquine and primaquine users, *C*_*T Q*_ = $47.89, *C*_*PQ*_ = $3.10, and *C*_*G*6*PD*_ = $7.76 are per-patient treatment and screening costs, *N*_*prevented*_ denotes the number of prevented relapse cases, *C*_*medical*_ is the cost per malaria case averted. The subscripts *Si* and *SA* denote intervention scenario *i* and the baseline (no-tafenoquine) scenario, respectively, where *i* ∈ {1, 2, 3, 4} represents the four deployment strategies.

Table 4 shows the economic metrics across all scenarios. Compared to baseline scenario A (SA), total costs increase from $2,741.07 to $9,925.88 (scenario E), while total benefits (TB) rise from $0 to $43,422.04. BCR values improve monotonically from Scenario A (SA, baseline) to 4.4 (scenario E), with NPV increasing from -$2,741.07 to $33,496.2. Incremental analysis relative to baseline reveals that scenario E provides IBCR = 6.04, representing the marginal economic efficiency of maximum tafenoquine deployment.

**Table 4:**
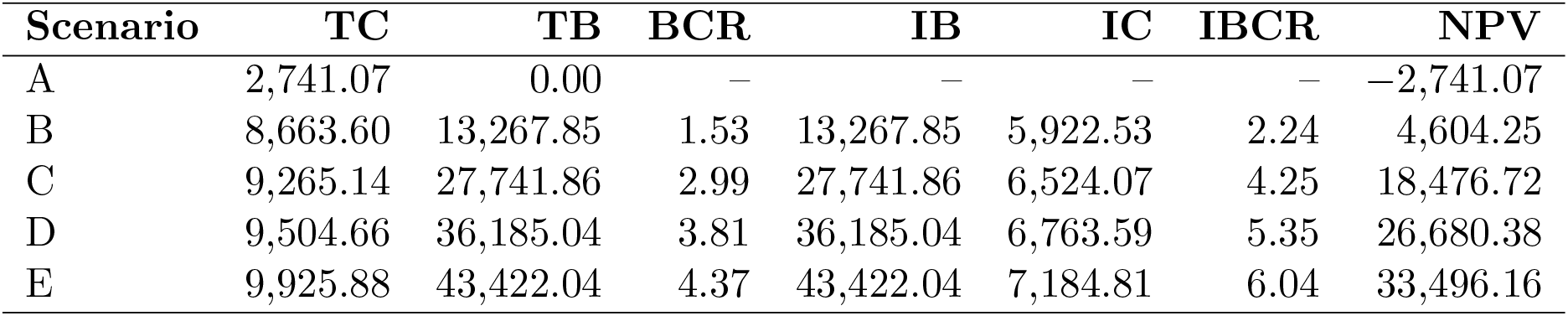
Economic evaluation metrics across tafenoquine deployment scenarios. Scenario A represents our baseline simulation where we use primaquine only. TC = Total Cost, TB = Total Benefit, BCR = Benefit-Cost Ratio, IB = Incremental Benefit, IC = Incremental Cost, IBCR = Incremental Benefit-Cost Ratio, NPV = Net Present Value. All monetary values in 2024 USD.

| Scenario | TC | TB | BCR | IB | IC | IBCR | NPV |
| --- | --- | --- | --- | --- | --- | --- | --- |
| A | 2,741.07 | 0.00 | – | – | – | – | –2,741.07 |
| B | 8,663.60 | 13,267.85 | 1.53 | 13,267.85 | 5,922.53 | 2.24 | 4,604.25 |
| C | 9,265.14 | 27,741.86 | 2.99 | 27,741.86 | 6,524.07 | 4.25 | 18,476.72 |
| D | 9,504.66 | 36,185.04 | 3.81 | 36,185.04 | 6,763.59 | 5.35 | 26,680.38 |
| E | 9,925.88 | 43,422.04 | 4.37 | 43,422.04 | 7,184.81 | 6.04 | 33,496.16 |

No discounting was applied given the one-year intervention timeframe, consistent with standard practice for short-term economic evaluations. The results demonstrate that despite 15-fold higher pharmaceutical costs, relapse prevention benefits justify tafenoquine investment across all coverage levels, with optimal returns achieved under unrestricted supply conditions.

## 7. Discussion and conclusion

This study introduces a novel, integrative modeling framework that optimizes malaria intervention strategies under climate-sensitive transmission dynamics. By coupling a relapse- and climate-aware epidemiological model with a SI assessment and a metaheuristic optimization approach, the framework provides a rigorous basis for designing adaptive control policies in elimination settings. Unlike previous studies that rely on simplified or locally tuned control formulations, our approach explicitly links model identifiability, uncertainty, and real-world intervention constraints. The Improved Multi-Operator Differential Evolution (IMODE) algorithm proved highly effective in navigating the 48-dimensional, non-convex decision space, achieving robust solutions across multiple climate and policy scenarios. This represents one of the first applications of a global evolutionary algorithm to optimize multi-intervention malaria control strategies under realistic operational and stockpile constraints. The mathematical model was specifically developed and tailored to the Korean malaria context through rigorous identifiability analysis and parameter estimation from surveillance data. The model demonstrated well-calibrated predictions and well-validated performance through independent data testing and comprehensive sensitivity analyses before the analysis of interventions.

Our primary findings demonstrate substantial benefits from tafenoquine deployment strategies. Complete tafenoquine substitution achieved 80% reduction in relapse cases and 16.6% reduction in total malaria cases compared to primaquine-only treatment. These epidemiological gains translate into strong economic returns, with incremental benefit-cost ratios reaching 6.04 despite 15-fold higher pharmaceutical costs. The consistent cost-effectiveness across coverage levels supports policy recommendations for expanding tafenoquine availability in populations with low G6PD deficiency rates.

Climate scenario analysis reveals the critical importance of adaptive intervention planning. The optimization results demonstrate that higher-transmission scenarios necessitate earlier intervention initiation and more sustained implementation, with optimal strategies exhibiting scenario-specific timing and intensity requirements (detailed in Appendix C). Under Seoul’s analyzed climate conditions, optimal adult mosquito control demonstrates dual-phase implementation targeting early spring overwintering populations (March-April) and peak summer transmission (July-September), while contact reduction achieves maximum effectiveness during mid-summer (June-August). This climate-responsive optimization addresses critical gaps in conventional static strategies that fail to account for transmission seasonality driven by temperature-dependent mosquito dynamics and relapse periodicity.

The case study conducted in Seoul demonstrates the practical applicability of the proposed framework to urban malaria control efforts. However, the current analysis excludes neighboring endemic provinces (Gyeonggi, Incheon) where substantial cross-regional transmission occurs. Extension to these regions would strengthen applicability to the broader Korean transmission network. Additionally, the deterministic framework does not incorporate stochastic environmental fluctuations or outbreak events, which may affect intervention timing in practice.

Future research could extend this framework to broader geographic contexts, including transboundary transmission dynamics with North Korea and other *P. vivax* endemic regions. Multi-regional modeling could capture spatial heterogeneity and cross-border epidemiological interactions that significantly influence local transmission patterns. Additionally, incorporating vector resistance patterns and seasonal migration effects would enhance model realism.

The demonstrated integration of climate-adaptive epidemiological modeling with optimization techniques provides a replicable methodology for evidence-based intervention planning. This case study establishes a foundation for developing region-specific malaria control strategies that account for local epidemiological, climatic, and economic conditions. The adaptable structure of the framework enables its application across diverse endemic settings, supporting global malaria elimination efforts through tailored, mathematically optimized intervention strategies.

## Data Availability

All data produced are available online at

https://www.kdca.go.kr/

https://data.kma.go.kr

## CRediT author statement

**Jiwon Han**: Conceptualization, Methodology, Software, Formal analysis, Investigation, Validation, Data curation, Visualization, Writing – original draft. **Gerardo Chowell**: Methodology, Validation, Writing – review & editing. **Eunok Jung**: Conceptualization, Methodology, Resources, Writing – review & editing, Supervision, Project administration, Funding acquisition.

## Data Availability

The epidemiological data that support the findings of this study are available from the Korea Disease Control and Prevention Agency (KDCA) at https://www.kdca.go.kr/. Climate data were obtained from the Korea Meteorological Administration at https://data.kma.go.kr.

## Appendix A Seasonal reproduction number (*R*_*s*_)

**Theorem 1** (Basic Reproduction Number). *For the P. vivax malaria transmission model given by equations (1), the basic reproduction number R*_*s*_ *is the spectral radius of the next-generation matrix FV* ^−1^.

*Proof*. We apply the next-generation matrix method [48]. Let **x** =(*E*_*V*_, *I*_*V*_, *E*_*HS*_, *E*_*HL*_, *I*_*H*_, *L*_*HS*_, *L*_*HL*_, *D*_*H*_ )^*T*^ represent the infected compartments. At the diseasefree equilibrium, all infected compartments are zero, and we have 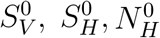 as the susceptible populations.

The matrices *F* (new infections) and *V* (transitions) are given by:

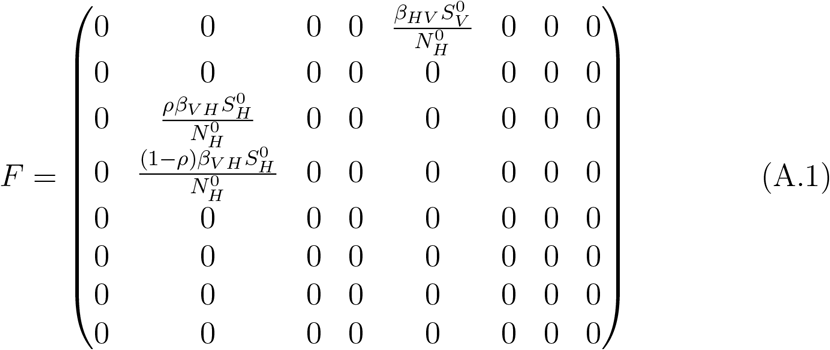

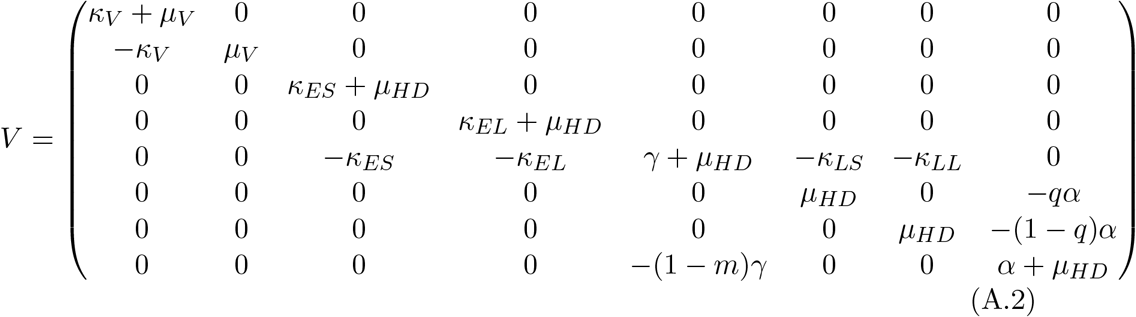

The next-generation matrix *FV* ^−1^ is computed as follows:

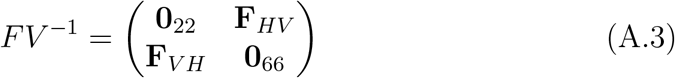

where the human-to-vector transmission block **F**_*HV*_ ∈ *ℝ*^2×6^ has transpose:

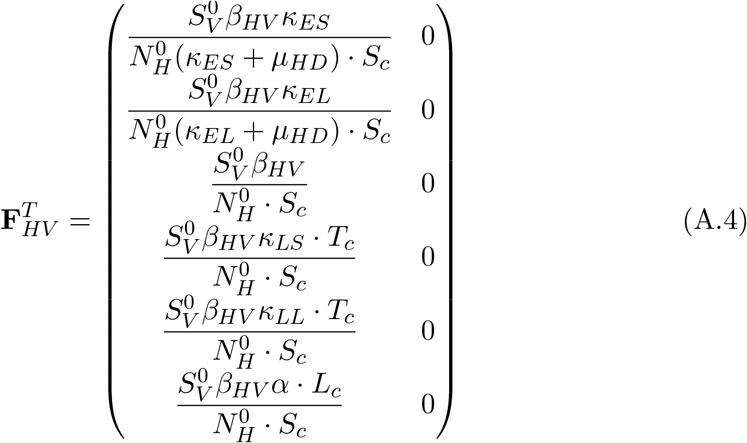

and the vector-to-human transmission block **F**_*V H*_ ∈ ℝ^6×2^ is:

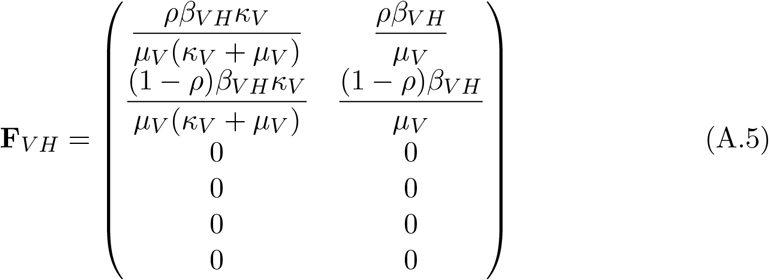

where the composite parameters are defined as:

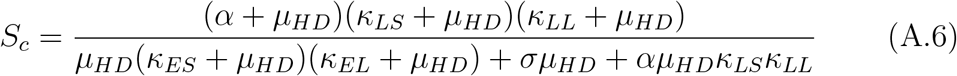

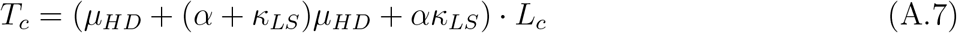

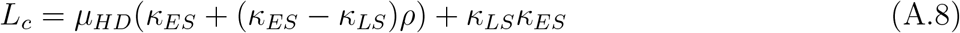

Here, *S*_*c*_ represents the survival composite capturing mortality and recovery rates, *T*_*c*_ represents the transition composite for inter-compartment flows, and *L*_*c*_ represents the latency composite for incubation periods.

The basic reproduction number *R*_*s*_ is obtained as the spectral radius of *FV* ^−1^, which simplifies to:

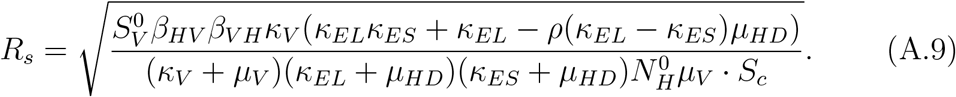

*R*_*s*_ represents the expected number of secondary infections produced by a single infectious individual in a completely susceptible population, accounting for P. vivax-specific features including relapse dynamics and seasonal mosquito population variations.

## Appendix B Parameter uncertainty quantification

We employed parametric bootstrap with *B* = 10, 000 replicates to quantify uncertainty in the estimated transmission parameters *β*_*HV*_ and *β*_*V H*_ . Out of 10,000 iterations, 9871 converged successfully and were retained for analysis.

Table B.5 presents the bootstrap parameter estimates. The coefficients of variation (CV = S.D./Mean × 100%) are 13.8% and 14.2% for *β*_*HV*_ and *β*_*V H*_ respectively, indicating moderate relative uncertainty. The bootstrap mean closely matches the point estimate (within 0.2% for both parameters), confirming the reliability of the original estimates. Confidence intervals exclude zero with substantial margin, demonstrating statistically significant transmission in both directions.

**Table B.5:** Bootstrap parameter estimates from 1,000 replicates. Parenthetical percentages show deviation of bootstrap mean from point estimate. Both parameters demonstrate moderate uncertainty (CV *<* 15%) with confidence intervals excluding zero.

| Parameter | Estimate | Bootstrap Mean | S.D. | 95% CI | CV (%) |
| --- | --- | --- | --- | --- | --- |
| $\beta_{HV}$ | 0.0452 | 0.0453 | $6.23 \times 10^{-3}$ | (0.0336, 0.0578) | 13.8 |
| $\beta_{VH}$ | 0.4996 | 0.4987 | $7.08 \times 10^{-2}$ | (0.3646, 0.6401) | 14.2 |

**Table D.6:**
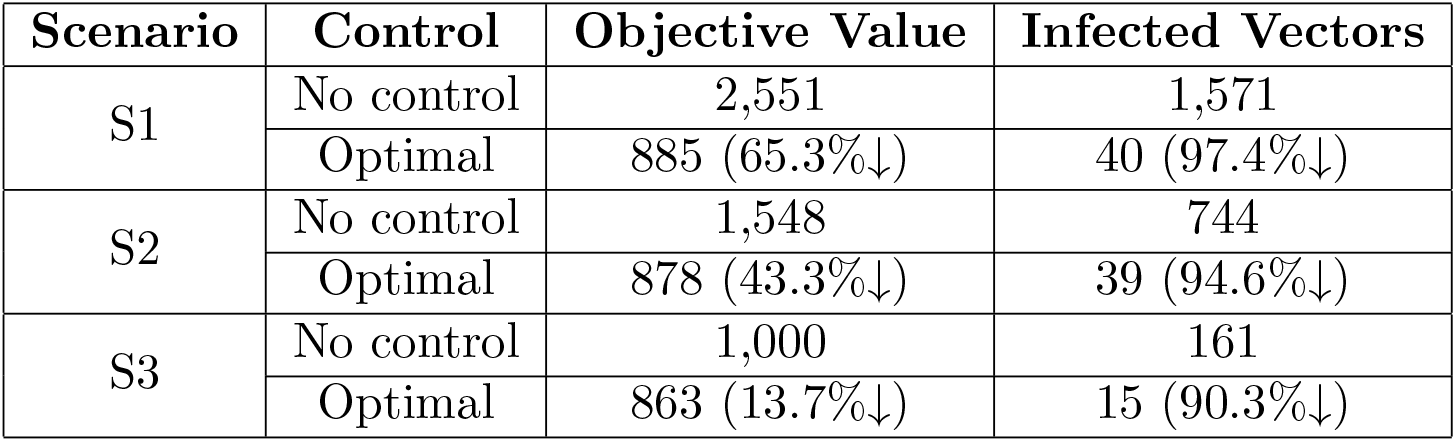
Effectiveness of optimal non-pharmaceutical interventions across climate scenarios. Parenthetical values show percentage reductions from baseline (no-control) conditions. Higher baseline transmission scenarios (S1) achieve greater relative reductions, while vector control remains highly effective (*>*90%) across all climate conditions.

| Scenario | Control | Objective Value | Infected Vectors |
| --- | --- | --- | --- |
| S1 | No control | 2,551 | 1,571 |
|  | Optimal | 885 (65.3%↓) | 40 (97.4%↓) |
| S2 | No control | 1,548 | 744 |
|  | Optimal | 878 (43.3%↓) | 39 (94.6%↓) |
| S3 | No control | 1,000 | 161 |
|  | Optimal | 863 (13.7%↓) | 15 (90.3%↓) |

S1, peak implementation occurs during July-August with maximum intensity (0.5), while scenario S2 requires moderate intervention (intensity $\sim 0.2$ ) during the same period. Notably, even the coolest scenario S3 necessitates baseline contact reduction measures (intensity $\sim 0.1$ ), indicating that this intervention remains essential across all climate conditions to maintain transmission control.

The substantially lower 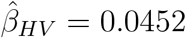 compared to 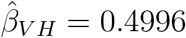 reflects the biological asymmetry in P. vivax transmission: low parasitemia in human infections reduces human-to-vector transmission efficiency, while prolonged mosquito infectious periods and repeated blood-feeding increases vector-to-human transmission. The bootstrap standard errors (Table B.5) quantify sampling uncertainty conditional on the model structure and transmission rates.

Figure B.8 shows the marginal distributions of bootstrap parameter estimates. Both distributions are approximately normal, consistent with asymptotic theory for nonlinear least-squares estimators. The relatively moderate spread indicates that the 35-week surveillance period (91 total cases) provides reasonable information for constraining these parameters. The bootstrap ensemble was used to generate prediction confidence intervals in Figure 2, where observed cases consistently fall within 95% bootstrap confidence bands, validating both parameter estimates and model structure.

**Figure B.8:**
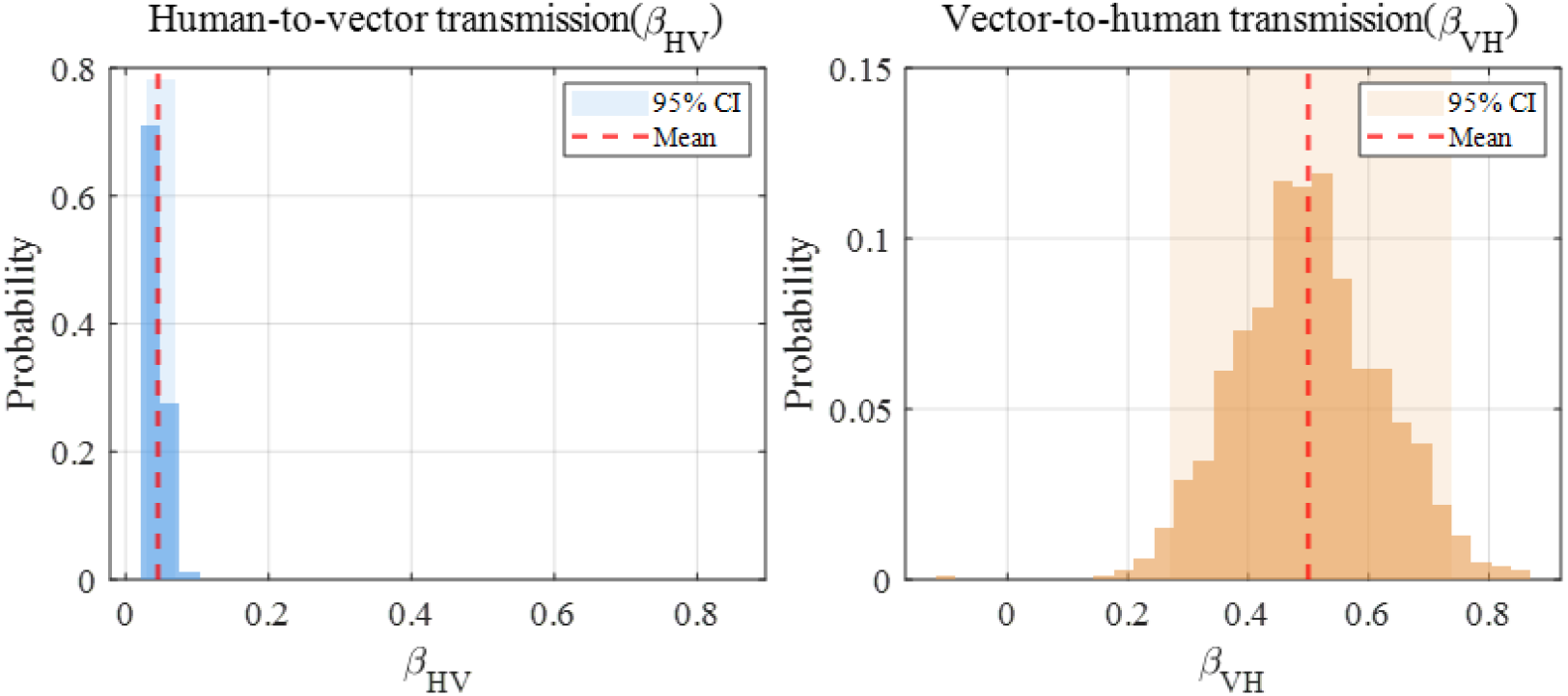
Bootstrap distributions from B = 10,000 replicates showing (A) *β*_*HV*_ (human-to-vector transmission) and (B) *β*_*V H*_ (vector-to-human transmission). Red dashed lines indicate point estimates 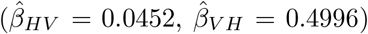. Shaded regions represent 95% confidence intervals. Both distributions exhibit approximate normality, supporting asymptotic confidence intervals. The lower magnitude and tighter distribution of *β*_*HV*_ reflect reduced transmission efficiency from low-parasitemia P. vivax infections.

## Appendix C Correlation methods

Figure C.9 presents correlation heatmaps summarizing the relationships between climate variables and mosquito infection burden across all scenarios.

**Figure C.9:**
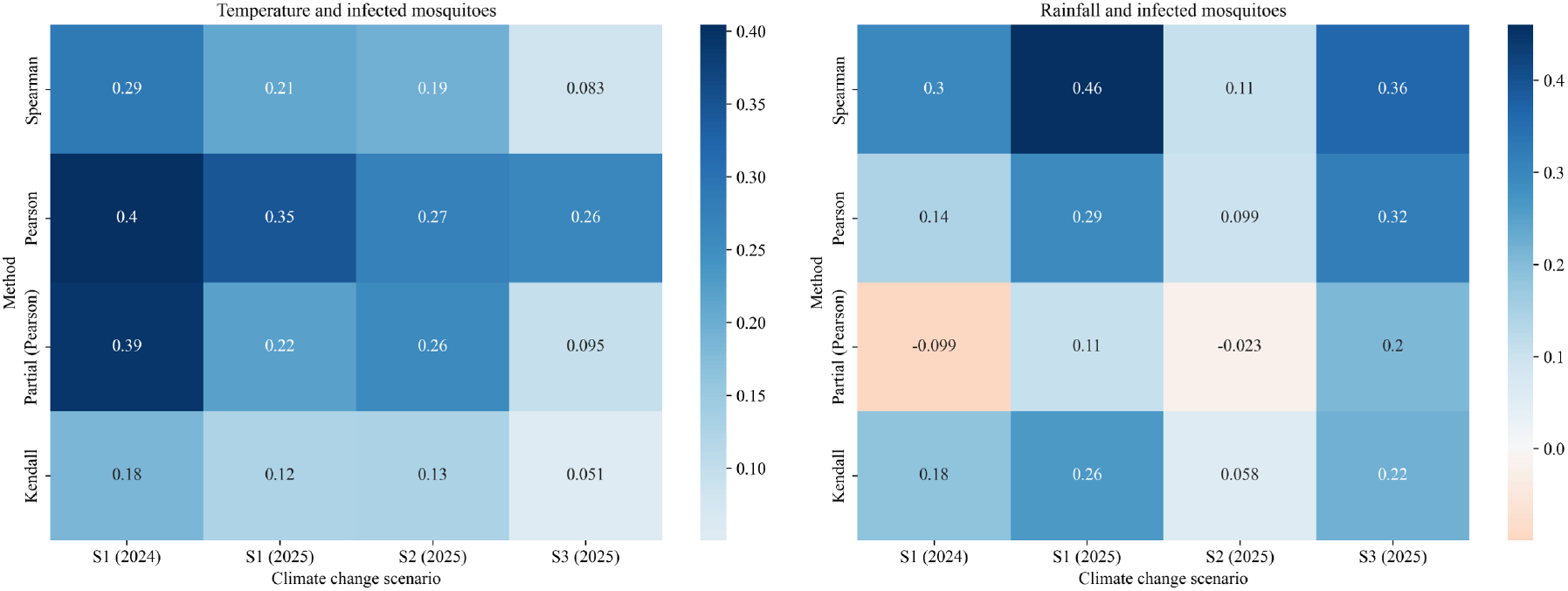
Correlation heatmaps between climate variables and cumulative infected mosquitoes across climate scenarios. Left panel shows temperature correlations, right panel shows precipitation correlations. Values represent correlation coefficients using four methods: Spearman, Pearson, Partial Pearson (controlling for the other climate variable), and Kendall’s tau across scenarios S1-S3 for years 2024-2025.

Four correlation methods were employed to capture different aspects of the climate-mosquito relationship [49, 50, 51, 52]:

- **Pearson correlation** measures linear relationships between continuous variables, assuming normal distributions [49] .
- **Spearman correlation** assesses monotonic relationships using rank-based statistics, robust to non-normal distributions and outliers [50].
- **Kendall’s tau** provides an alternative rank-based measure with better small-sample properties and resistance to tied values [51].
- **Partial Pearson correlation** evaluates the linear association between two variables while controlling for the effect of a third variable, allowing isolation of direct relationships [52].

### Appendix C.1. Temperature correlations

Temperature demonstrates consistently positive correlations with infected mosquito burden across all methods and scenarios. Temperature shows consistently significant correlations (p *<* 0.001 for all scenarios). Pearson correlations range from 0.4042 (S1, 2024) to 0.4249 (S2, 2025), with all relationships highly significant (p *<* 0.001). Spearman correlations show similar patterns (0.2967 to 0.4385), indicating robust monotonic relationships. Kendall’s tau values (0.1841 to 0.3075) confirm the positive association, while Partial Pearson correlations (0.3929 to 0.4242) demonstrate that temperature effects remain significant after controlling for precipitation.

### Appendix C.2. Precipitation correlations

Precipitation shows more variable correlations across scenarios, while precipitation correlations vary in significance across climate scenarios (p-values ranging from 0.0011 to 0.1116). Positive associations are strongest in S1 (2025) and S3 (2025) scenarios (Pearson r = 0.2931 and 0.3174 respectively, p *<* 0.001), while S2 (2025) shows weaker correlation (r = 0.0999, p = 0.1116). Spearman correlations remain consistently positive and significant except for S2 (2025). Notably, Partial Pearson correlations become negative in some scenarios (S1 2024: r = -0.0992, S2 2025: r = -0.0234), suggesting complex interactions between temperature and precipitation effects.

### Appendix C.3. Implications

The stronger and more consistent temperature correlations indicate that thermal conditions are the primary driver of mosquito infection dynamics, while precipitation effects are more scenario-dependent and may be moderated by temperature interactions. These findings support the temperature-dependent parameterization approach used in the main model.

## Appendix D Extended results for optimal NPIs

### Appendix D.1. Scenario-based control effectiveness

The effectiveness of intervention strategies varies significantly across climate scenarios. Under baseline conditions without control, scenario S1 shows the highest transmission burden with 2,551 cumulative infections and 1,571 infected vectors, reflecting the most favorable conditions for *P. vivax* transmission. Scenarios S2 and S3 demonstrate progressively lower baseline transmission (1,548 and 1,000 infections respectively), indicating reduced mosquito activity under altered climate conditions.

When optimal non-pharmaceutical interventions are applied, all scenarios achieve substantial reductions. S1 shows 65.3% reduction in infections (885 cases) and 97.4% reduction in infected vectors. S2 and S3 demonstrate similar control efficacy with 43.3% and 13.7% reductions in infections respectively.

The higher absolute effectiveness in S1 reflects both the greater baseline burden and more intensive optimal control strategies required under hightransmission conditions.

### Appendix D.2. Optimal intervention timing across climate scenarios

The temporal patterns of optimal non-pharmaceutical interventions reveal scenario-specific adaptation to climatic conditions (Figure D.11).

**Figure D.10:**
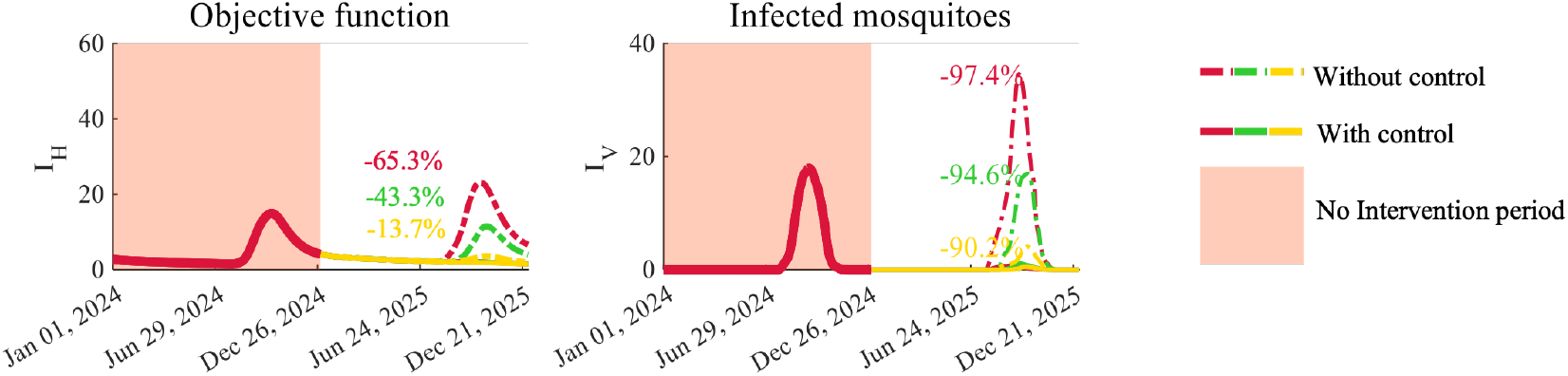
Control effectiveness across climate scenarios showing objective function reduction and infected mosquito dynamics with quantitative reduction percentages.

**Figure D.11:**
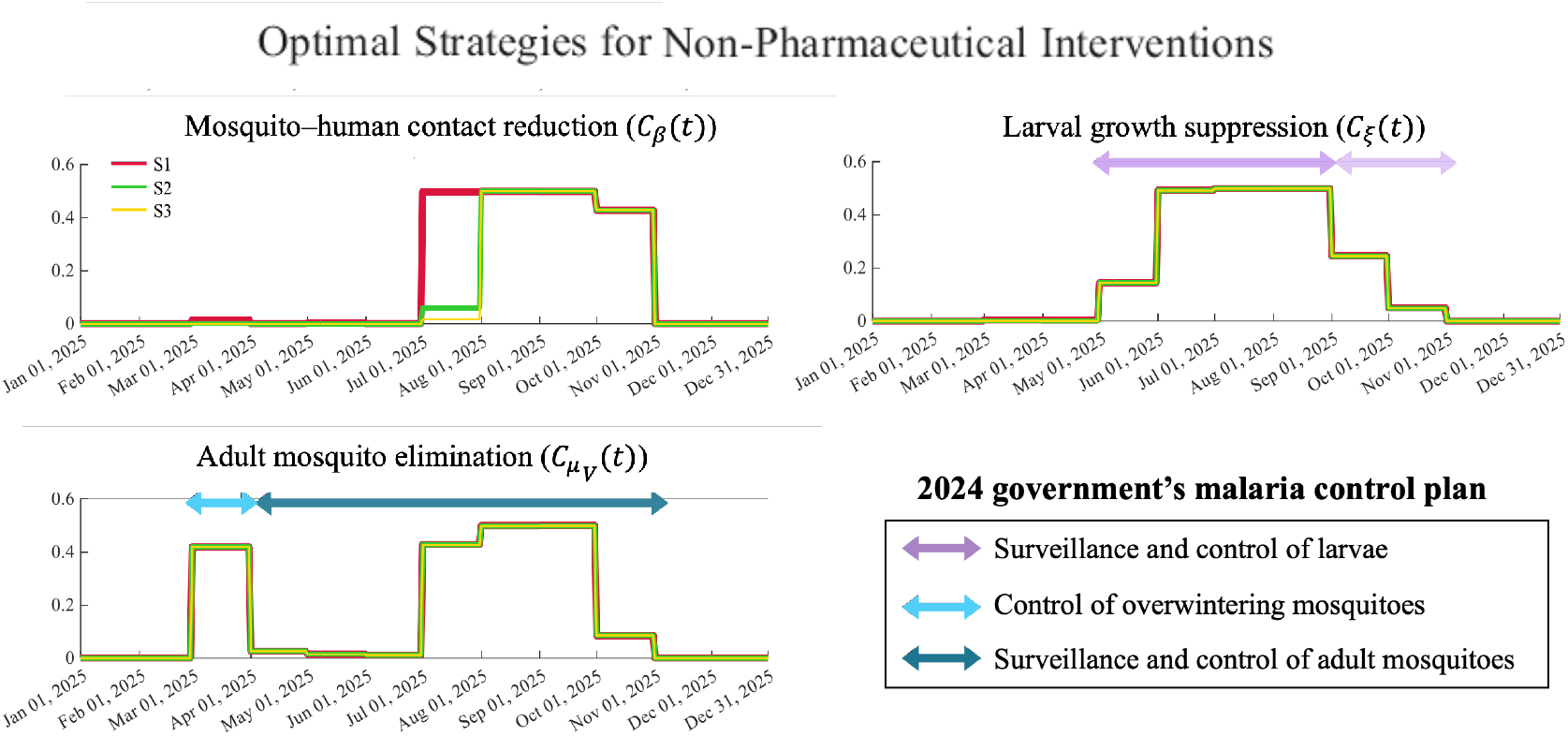
Optimal temporal patterns for non-pharmaceutical interventions across climate scenarios S1-S3, aligned with 2024 government malaria control plan.

#### Mosquito-human contact reduction

(*C*_*β*_(*t*)) demonstrates the strongest climate dependency among all interventions. Under the warmest scenario S1, peak implementation occurs during July-August with maximum intensity (0.5), while scenario S2 requires moderate intervention (intensity ∼0.2) during the same period. Notably, even the coolest scenario S3 necessitates baseline contact reduction measures (intensity ∼0.1), indicating that this intervention remains essential across all climate conditions to maintain transmission control.

#### Larval growth suppression

(*C*_*ξ*_(*t*)) shows remarkably consistent timing across all scenarios, with optimal implementation concentrated during June-August at maximum intensity (0.5) regardless of climate conditions. This uniformity suggests that larval control effectiveness is primarily determined by mosquito breeding season rather than climate-driven transmission intensity. While government guidelines recommend larvicidal activities from May to September with optional extension through November when conditions warrant, our optimization results indicate that sustained low-intensity implementation (0.1-0.2) during September-November provides additional transmission reduction benefits at minimal cost, supporting the government’s flexible extension policy.

#### Adult mosquito elimination

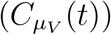 demonstrates dual-phase implementation patterns that align closely with government malaria control strategies. All scenarios exhibit early spring intervention (March-April) targeting overwintering mosquito populations, followed by intensive summer control (July-September) during peak transmission periods. Notably, optimization results indicate that low-intensity control (0.1-0.2) during November remains beneficial for preventing late-season transmission, consistent with government recommendations for extended surveillance and control activities. The dual-phase timing (spring and summer-autumn) directly corresponds to the government’s 2024 operational plan, which designates February-April for overwintering mosquito control (blue arrow) and June-September for active season interventions (dark blue arrow).

These optimization-derived strategies demonstrate substantial concordance with the 2024 Korean government malaria control plan. These optimization-derived strategies were developed using the same epidemiological data and operational constraints that inform government planning, resulting in substantial concordance with the 2024 Korean government malaria control plan.

The government’s larval surveillance and control period (purple arrow: April-October with optional November extension) directly encompasses the optimal June-August intensive period plus the recommended low-intensity September-November activities identified by optimization. The early spring focus on overwintering mosquito elimination (blue arrow: February-April) matches the March-April optimal intervention window across all climate scenarios. Peak summer intervention timing (dark blue arrow: June-September) corresponds precisely to the July-September intensive control period derived from optimization. This concordance confirms that the optimization framework successfully captures operational realities and epidemiological dynamics underlying field-validated control practices, while additionally quantifying optimal intensity levels and climate-adaptive adjustments not explicitly specified in government guidelines.

## References

[1] White, N.J., Determinants of relapse periodicity in Plasmodium vivax malaria, Malaria Journal, vol. 10, 297, 2011.

[2] White, M.T., Karl, S., Battle, K.E., Hay, S.I., Mueller, I., Ghani, A.C., Modelling the contribution of the hypnozoite reservoir to Plasmodium vivax transmission, eLife, vol. 3, e04692, 2014.

[3] Caminade, C., Kovats, S., Rocklov, J., Tompkins, A.M., Morse, A.P., Colón-González, F.J., Stenlund, H., Martens, P., Lloyd, S.J., Impact of climate change on global malaria distribution, Proceedings of the National Academy of Sciences, vol. 111, no. 9, pp. 3286–3291, 2014.

[4] Ryan, S.J., Carlson, C.J., Mordecai, E.A., Johnson, L.R., Global expansion and redistribution of Aedes-borne virus transmission risk with climate change, PLoS Neglected Tropical Diseases, vol. 13, no. 3, e0007213, 2019.

[5] Anderson, R.M., May, R.M., Infectious Diseases of Humans: Dynamics and Control, Oxford University Press, Oxford, 1991.

[6] Griffin, J.T., Hollingsworth, T.D., Okell, L.C., Churcher, T.S., White, M., Hinsley, W., Bousema, T., Drakeley, C.J., Ferguson, N.M., Basánez, M.G., Ghani, A.C., Reducing Plasmodium falciparum malaria transmission in Africa: a model-based evaluation of intervention strategies, PLOS Medicine, vol. 7, no. 8, e1000324, 2010. “‘

[7] Lenhart, S., Workman, J.T., Optimal Control Applied to Biological Models, Chapman and Hall/CRC, 2007.

[8] Adekunle, A.I., Pinkevych, M., McGready, R., Luxemburger, C., White, L.J., Nosten, F., Cromer, D., Davenport, M.P., Modeling the Dynamics of Plasmodium vivax Infection and Hypnozoite Reactivation In Vivo, PLOS Neglected Tropical Diseases, vol. 9, no. 3, e0003595, 2015.

[9] Villena, O.C., Ryan, S.J., Murdock, C.C., Johnson, L.R., Temperature impacts the environmental suitability for malaria transmission by Anopheles gambiae and Anopheles stephensi, Ecology, vol. 103, no. 8, e3685, 2022.

[10] Shocket, M.S., Bernhardt, J.R., Miazgowicz, K.L., Orakzai, A., Savage, V.M., Hall, R.J., Ryan, S.J., Murdock, C.C., Mean daily temperatures predict the thermal limits of malaria transmission better than hourly rate summation, Nature Communications, vol. 16, Article 3441, 2025.

[11] Kim, S., Byun, J.H., Park, A., Jung, I.H., A mathematical model for assessing the effectiveness of controlling relapse in Plasmodium vivax malaria endemic in the Republic of Korea, PLOS ONE, vol. 15, no. 1, e0227919, 2020.

[12] Bahk, Y.Y., Lee, H.W., Na, B.K., Kim, J., Jin, K., Hong, Y.S., Kim, T.S., Epidemiological Characteristics of Re-emerging Vivax Malaria in the Republic of Korea (1993–2017), The Korean Journal of Parasitology, vol. 56, no. 6, pp. 531–543, 2018.

[13] Price, D.J., Nekkab, N., Monteiro, W.M., Villela, D.A.M., Simpson, J.A., Lacerda, M.V.G., et al., Tafenoquine following G6PD screening versus primaquine for the treatment of vivax malaria in Brazil: A costeffectiveness analysis using a transmission model, PLOS Medicine, vol. 21, no. 1, e1004255, 2024.

[14] Suh, J., Kim, J.H., Kim, J.D., Kim, C., Choi, J.Y., Lee, J., Yeom, J.S., Cost-Benefit Analysis of Tafenoquine for Radical Cure of Plasmodium vivax Malaria in Korea, Journal of Korean Medical Science, vol. 37, no. 27, e212, 2022.

[15] Chowell, G., Dahal, S., Liyanage, Y.R., Tariq, A., Tuncer, N., Structural identifiability analysis of epidemic models based on differential equations: a tutorial-based primer, Journal of Mathematical Biology, vol. 87, no. 6, 79, 2023.

[16] Dankwa, E.A., Brouwer, A.F., Donnelly, C.A., Structural identifiability of compartmental models for infectious disease transmission is influenced by data type, Epidemics, vol. 41, 100643, 2022.

[17] Lacerda, M.V., Llanos-Cuentas, A., Krudsood, S., Lon, C., Saunders, D.L., Mohammed, R., Yilma, D., Batista Pereira, D., Espino, F.E., Xie, L.H., Millán, S.B., Hamed, K., Maher, S.P., Moehrle, J.J., Chalon, S., Single-dose tafenoquine to prevent relapse of Plasmodium vivax malaria, New England Journal of Medicine, vol. 380, no. 3, pp. 215–228, 2019.

[18] Anwar, M.N., Hickson, R.I., McCaw, J.M., Quam, M.B., Morshed, M.S., Islam, A., Mathematical models of Plasmodium vivax transmission: A scoping review, PLoS Computational Biology, vol. 20, no. 3, e1011931, 2024.

[19] Beck-Johnson, L.M., Nelson, W.A., Paaijmans, K.P., Read, A.F., Thomas, M.B., Bjørnstad, O.N., The effect of temperature on Anopheles mosquito population dynamics and the potential for malaria transmission, PLoS One, vol. 8, no. 11, e79276, 2013.

[20] Abdelrazec, A., Gumel, A.B., Mathematical assessment of the role of temperature and rainfall on mosquito population dynamics, Journal of Mathematical Biology, vol. 74, no. 6, pp. 1351–1395, 2017.

[21] Kim JE, Choi Y, Lee CH, Effects of climate change on Plasmodium vivax malaria transmission dynamics: a mathematical modeling approach, Applied Mathematics and Computation, 347:616–630, 2019.

[22] Paaijmans KP, Cator LJ, Thomas MB, Temperature-dependent pre-bloodmeal period and temperature-driven asynchrony between parasite development and mosquito biting rate reduce malaria transmission intensity, PLoS ONE, 8(3):e55777, 2013.

[23] Chitnis N, Cushing JM, Hyman J, Bifurcation analysis of a mathematical model for malaria transmission, SIAM Journal on Applied Mathematics, 67(1):24–45, 2006.

[24] Okuneye K, Gumel AB, Analysis of a temperature- and rainfall-dependent model for malaria transmission dynamics, Mathematical Biosciences, 287:72–92, 2017.

[25] Mandal S, Sarkar RR, Sinha S, Mathematical models of malaria–a review, Malaria Journal, 10(1):202, 2011.

[26] Korea National Statistical Office (KOSIS), Future population projection data, https://www.korea.kr/briefing/policyBriefingView.do?newsId=156605259, 2023, Accessed: December 2024.

[27] Kim SJ, Kim SH, Jo SN, Gwack J, Youn SK, Jang JY, The Long and Short Incubation Periods of Plasmodium vivax Malaria in Korea: The Characteristics and Relating Factors, Infection & Chemotherapy, 45(2):184–193, 2013.

[28] Nah K, Kim Y, Lee JM, The dilution effect of the domestic animal population on the transmission of P. vivax malaria, Journal of Theoretical Biology, 266(2):299–306, 2010.

[29] Kwak YG, Lee HK, Kim M, Um TH, Cho CR, Clinical characteristics of vivax malaria and analysis of recurred patients, Infection & Chemotherapy, 45(1):69–75, 2013.

[30] White NJ, Determinants of relapse periodicity in Plasmodium vivax malaria, Malaria Journal, 10:297, 2011.

[31] Korea Centers for Disease Control and Prevention, Malaria management guidelines 2018, http://www.kdca.go.kr/npt/biz/npp/portal/nppPblctDtaView.do?pblctDtaSeAt=8&pblctDtaSn=883, 2018, Accessed: June 2021.

[32] Chamchod F, Beier JC, Modeling Plasmodium vivax: relapses, treatment, seasonality, and G6PD deficiency, Journal of Theoretical Biology, 316:25–34, 2013.

[33] Llanos-Cuentas A, Lacerda MV, Rueangweerayut R, et al., Tafenoquine plus chloroquine for the treatment and relapse prevention of Plasmodium vivax malaria (DETECTIVE): a multicentre, double-blind, randomised, phase 2b dose-selection study, Lancet, 383(9922):1049–1058, 2014.

[34] Baird JK, Chloroquine resistance in Plasmodium vivax, Antimicrobial Agents and Chemotherapy, 48(11):4075–4083, 2004.

[35] Beck-Johnson LM, Nelson WA, Paaijmans KP, Read AF, Thomas MB, Bjørnstad ON, The effect of temperature on Anopheles mosquito population dynamics and the potential for malaria transmission, PLoS ONE, 8(11):e79276, 2013.

[36] Korea Meteorological Administration, Korea Climate Information Portal: Daily temperature and precipitation data, https://data.kma.go.kr/climate/, 2023, Accessed: December 2024.

[37] Dong R, Goodbrake C, Harrington H, Pogudin G, Differential Elimination for Dynamical Models via Projections with Applications to Structural Identifiability, SIAM Journal on Applied Algebra and Geometry, 7(1):194–235, 2023.

[38] Liyanage YR, Saucedo O, Tuncer N, Chowell G, A Tutorial on Structural Identifiability of Epidemic Models Using StructuralIdentifiability.jl, arXiv preprint arXiv:2505.10517, 2025.

[39] Dankwa EA, Brouwer AF, Donnelly CA, Structural identifiability of compartmental models for infectious disease transmission is influenced by data type, Epidemics, 41:100643, 2022.

[40] Korea Disease Control and Prevention Agency, Infectious Disease Portal: Malaria Surveillance Data for Seoul, https://www.kdca.go.kr/, 2023, Accessed: December 2024.

[41] Korea Meteorological Administration, Korea Climate Information Portal: Daily Temperature and Precipitation Data, https://data.kma.go.kr/cmmn/main.do, 2023, Accessed: December 2024.

[42] Efron B, Tibshirani RJ, An Introduction to the Bootstrap, Chapman and Hall/CRC, 1994.

[43] Marino S, Hogue IB, Ray CJ, Kirschner DE, A methodology for performing global uncertainty and sensitivity analysis in systems biology, Journal of Theoretical Biology, 254(1):178–196, 2008.

[44] McKay MD, Beckman RJ, Conover WJ, A comparison of three methods for selecting values of input variables in the analysis of output from a computer code, Technometrics, 21(2):239–245, 1979.

[45] Sobol IM, Global sensitivity indices for nonlinear mathematical models and their Monte Carlo estimates, Mathematics and Computers in Simulation, 55(1–3):271–280, 2001.

[46] Sallam KM, Elsayed SM, Chakrabortty RK, Ryan MJ, Improved Multi-operator Differential Evolution Algorithm for Solving Unconstrained Problems, In: 2020 IEEE Congress on Evolutionary Computation (CEC), IEEE, pp. 1–8, 2020.

[47] Korea Disease Control and Prevention Agency, 2024 Malaria Elimination Research Project and Operational Plan, Available at: https://eng.phwr.org/journal/view.html?pn=search&uid=703&vmd=Full, 2024.

[48] Van den Driessche P, Watmough J, Reproduction numbers and subthreshold endemic equilibria for compartmental models of disease transmission, Mathematical Biosciences, 180(1–2):29–48, 2002.

[49] Pearson K, Notes on regression and inheritance in the case of two parents, Proceedings of the Royal Society of London, 58:240–242, 1895.

[50] Spearman C, The proof and measurement of association between two things, American Journal of Psychology, 15(1):72–101, 1904.

[51] Kendall MG, A new measure of rank correlation, Biometrika, 30(1–2):81–93, 1938.

[52] Fisher RA, The distribution of the partial correlation coefficient, Metron, 3:329–332, 1924.

